# Defining severe shigellosis in the Enterics for Global Health study: A comparison of leading diarrhea severity definitions among children with *Shigella* diarrhea

**DOI:** 10.1101/2025.07.30.25331602

**Authors:** Olivia Lang Schultes, Aneeta Hotwani, Patricia B. Pavlinac, M. Jahangir Hossain, Dilruba Nasrin, Richard Omore, Farhana Khanam, Flywell Kawonga, Paul F. Garcia Bardales, Hannah E. Atlas, Alex O. Awuor, Henry Badji, Bakary Conteh, Jennifer Cornick, Erika L. Feutz, Sean R. Galagan, Ensa Gitteh, Eric R. Houpt, Junaid Iqbal, Sadia Islam, Furqan Kabir, Adama Mamby Keita, Jie Liu, Donnie Mategula, Chimwemwe Mhango, Md Parvej Mosharraf, Billy Ogwel, Caleb Okonji, Uduma Uma Onwuchekwa, S.M. Azadul Alam Raz, Lucero Romaina-Cachique, Francesca Schiaffino, Tackeshy Pinedo Vasquez, Mohammad Tahir Yousafzai, Milagritos D. Tapia, Sharon M. Tennant, Khuzwayo C. Jere, Margaret N. Kosek, Samba O. Sow, James A. Platts-Mills, Farah Naz Qamar, Elizabeth T. Rogawski McQuade, Karen L. Kotloff, John Benjamin Ochieng, Firdausi Qadri, the EFGH Consortium

## Abstract

**Background:** *Shigella* has high global burden and serious long-term outcomes among young children. Increasing antibiotic resistance makes *Shigella* vaccine development a critical priority. Choosing the optimal clinical efficacy endpoint for *Shigella* vaccine trials requires comparing existing diarrhea severity definitions among children with *Shigella*.

**Methods:** Six-to 35-month-old children presenting to a clinic with diarrhea were enrolled in Bangladesh, Kenya, Malawi, Mali, Pakistan, Peru, and The Gambia. Among children with *Shigella*, six dichotomized diarrhea severity definitions (GEMS moderate-to-severe diarrhea (MSD), Modified Vesikari Score (MVS), MVS +/− dysentery, Clark score, MAL-ED score, and *Shigella* mortality score) were examined against two outcomes: change in length/height-for-age z-score (ΔLAZ/HAZ) from enrollment to three-month follow-up and death or hospitalization within fourteen days of enrollment. We compared the performance of each severity score to predict outcomes using linear regression, measures of diagnostic accuracy, and Receiver Operating Characteristic (ROC) curves.

**Results:** Among 1,968 children with *Shigella*, moderate or severe diarrhea was negatively associated with ΔLAZ/HAZ by three definitions: MVS (−0.07 z-score, 95% CI −0.10 to −0.03), MVS +/− dysentery (−0.05, 95% CI −0.09 to −0.02), and MAL-ED score (−0.04, 95% CI −0.07 to 0.00). GEMS MSD (93.6%), MAL-ED (87.2%), MVS +/− dysentery (85.1%), and MVS (80.9%) had high sensitivity in predicting death or hospitalization, while *Shigella* mortality had the largest area under the curve (91.0).

**Conclusions:** MVS, MVS +/− dysentery, and the MAL-ED score identified adverse outcomes following *Shigella* diarrhea, indicating that they are viable options for a *Shigella* vaccine trial case definition.

**40-word summary:** We compared six diarrhea severity definitions among children 6-35 months with *Shigella*. Moderate-to-severe diarrhea as measured by Modified Vesikari Score (MVS), MVS +/− dysentery, and MAL-ED score was associated with reduced length/height-for-age z-score and hospitalization or mortality.

## Background

*Shigella* is a major cause of diarrhea among young children[1] causing approximately 60,000 deaths annually in children under five[2]. *Shigella* diarrhea also leads to an estimated 2 million cases of moderate-to-severe childhood stunting annually, and deaths caused by stunting increase the mortality burden of acute *Shigella* infections by 28%[2]. High disease burden, serious long-term consequences, and increasing resistance to common antibiotics support the need for making the development of *Shigella* vaccines an urgent global priority.

Several O-antigen vaccines targeting the most common *Shigella* serotypes (*S. flexneri* 1b, 2a, 3a, and 6, and *S. sonnei*) are under development[3–5]. Phase III licensure trials require a validated definition of clinical severity for the primary endpoint. Most licensed vaccines have demonstrated higher efficacy against more severe disease, including vaccines for rotavirus[6], cholera[7], dengue[8], and influenza[9], where licensure decisions were based on the demonstration of efficacy against more severe clinical presentation. This is expected to be true for *Shigella* as well; a human challenge study of the monovalent biconjugate *Shigella* vaccine, *Flexyn2a,* found 30% efficacy against mild disease and 72% efficacy against severe shigellosis[10]. From a public health and health system cost perspective, prevention of more severe disease is also a high priority.

Several diarrhea severity definitions could be proposed for a vaccine trial. The Global Enteric Multicenter Study moderate-to-severe diarrhea[1] (GEMS MSD) definition identified children in low- and middle-income countries (LMICs) with significantly higher frequency of episode-related death and stunting[1]. The *Shigella* mortality score was developed to distinguish children with *Shigella-* attributable MSD at high/low risk of death[11]. The modified Vesikari score (MVS) accounts for characteristics over the complete diarrhea episode[12] and predicts hospitalization among diarrhea cases[13]. The combination of MVS and visible blood in stools has also been posed as a severity-defining tool[14]. The Clark and MAL-ED scores are quantitative measures of diarrhea severity and were associated with hospitalization in the context of all-cause diarrhea in a community setting[15].

Choosing the optimal clinical severity definition for efficacy endpoints to be used in *Shigella* vaccine trials requires a head-to-head comparison of existing definitions in children with *Shigella*-attributable medically-attended diarrhea (MAD). With detailed clinical data, the use of both culture and quantitative real-time PCR (qPCR) for laboratory confirmation of *Shigella*, a large sample size[5,16,17], and longitudinal follow-up, the Enterics for Global Health (EFGH) study enabled a comprehensive evaluation of six diarrhea severity definitions. We established how well severity definitions predict *Shigella*-specific adverse outcomes such as linear growth faltering, hospitalization, and death-outcomes that a *Shigella* vaccine will aim to avert.

## Methods

The EFGH *Shigella* surveillance study took place at clinical sites in Bangladesh, Kenya, Malawi, Mali, Pakistan, Peru, and The Gambia and aimed to establish incidence rates of *Shigella* diarrhea[5]. We enrolled children aged 6-35 months presenting to healthcare facilities with diarrhea and followed them for three months. Recruitment and follow up occurred between June 2022 and December 2024. Clinical research forms and other study documentation were prepared centrally and approved by local institutional review boards prior to enrollment[5].

At enrollment, caregivers reported the child’s symptoms since episode onset and clinicians assessed temperature, signs of dehydration, and behavior[16]. Caregivers were given a paper diarrhea diary with visual cues and were trained to record daily information on the number of loose stools, dysentery, vomiting, and temperature for the following fourteen days. At week four and month three follow-up visits, caregivers returned the diary and answered questions about symptoms during the index episode since enrollment. Hospitalization information was abstracted from records and caregiver recall at follow-up visits, and deaths were identified through caregiver report at follow-up visits (methods Appendix). Trained study staff measured participant weight and length/height at enrollment and follow-up visits, with refresher trainings and standardization tests conducted every six months[16].

Three rectal swabs were taken at enrollment and were placed in a dry cryovial tube for qPCR testing, modified-buffer glycerol saline (mBGS), and Cary-Blair media, respectively. Participants were classified as having *Shigella*-attributable MAD if *Shigella* was identified via culture in either media type or by qPCR with a cycle threshold less than 29.5[17,18]. Whole stool was also collected and tested using qPCR in Bangladesh and The Gambia, and qPCR results from whole stool (using a cycle threshold cutoff of 29.8[17]) were used if rectal swab results were unavailable (n=194). Staff training and data quality plans ensured high data quality[19].

Among participants who returned the diarrhea diary (n=9,032, 95.3%), a record of all four severity indicators was complete for 90.9% of participants. Severity indicators were constructed using data from the diarrhea diary, enrollment, and follow-up visits; all children had information from at least one source for each severity indicator. Length/Height-for-age z-scores (LAZ/HAZ) were derived per World Health Organization guidelines[20], and the difference from enrollment to the three-month follow-up visit (ΔLAZ/HAZ) was calculated for each participant with both measurements. LAZ/HAZ scores <–6 or >6 were considered implausible and excluded[21], as were length/height increases of >6 cm or losses of > 1.5cm from enrollment to three-month follow-up.

Severity indicator (dehydration, duration ≥6 days, dysentery, fever, and vomiting) frequencies are presented among *Shigella*-attributable MAD and all MAD. We classified each child according to the following severity definitions: MVS[12], moderate or severe diarrhea by MVS or dysentery (MVS +/− dysentery)[14], GEMS MSD[1], *Shigella* mortality score[11], Clark score[22], and MAL-ED score[15] (methods Appendix, Table A1). The distribution of severity definition categories (mild vs. moderate or severe) was presented among *Shigella*-attributable MAD overall and by site, age group, and culture positivity.

Among *Shigella*-attributable MAD and all MAD, we estimated the association between each dichotomized severity score and the continuous outcome of ΔLAZ/HAZ using linear regression. Models were stratified by receipt of guideline-recommended antibiotics for shigellosis[23] (azithromycin, ciprofloxacin, ceftriaxone, or pivmecillinam) since treatment may affect the relationship between shigellosis severity and adverse outcomes[24]. Generalized estimating equations using robust standard errors were used to account for clustering of children who were enrolled for more than one diarrhea episode, and models were adjusted for enrollment LAZ/HAZ, site, and days from enrollment to three-month follow-up visit.

We determined the diagnostic accuracy of each dichotomized score in predicting hospitalization or death within fourteen days of enrollment, separately among *Shigella*-attributable MAD and all MAD and stratified by antibiotic treatment. Sensitivity, specificity, accuracy, positive predictive value (PPV), and negative predictive value (NPV) were calculated. We calculated bootstrapped 95% confidence intervals (95% CIs) for each measure stratified by site and clustered by unique child. We performed Receiver Operating Characteristics (ROC) analysis to compare the performance of continuous severity scores on predicting the outcome of hospitalization or mortality among children with *Shigella*-attributable MAD.

Analyses were conducted using R Statistical Software (v4.3; R Core Team 2024).

## Results

Among 1,968 children with *Shigella*-attributable MAD, 48.6% were female, 19.6% were aged 6-11 months, 55.0% were 12-23 months, and 25.5% were 24-35 months (Table 1). Overall, 21.2% of children had moderate or severe wasting at enrollment, 28.0% were stunted, and 28.0% were moderately or severely underweight. Diarrhea severity indicators varied across sites. The proportion of children with some or severe dehydration was high in Peru (85.9%) and Kenya (56.6%), but under 20% in other sites. When examining dehydration signs across sites, child’s condition (irritable/lethargic) was the most reported sign (40.3%), followed by ability to drink (thirsty/unable to drink, 25.5%), sunken eyes (17.6%), and slow/very slow skin pinch (11.9%). Similar patterns were observed among all children with MAD (Table A2).

**Table 1.** Demographic characteristics and diarrhea severity indicators of participants with *Shigella*-attributable diarrhea by site.

| Characteristic | Bangladesh<br>N = 388 | Kenya<br>N = 175 | Malawi<br>N = 173 | Mali<br>N = 240 | Pakistan<br>N = 368 | Peru<br>N = 227 | The Gambia<br>N = 397 | Overall<br>N = 1,968 |
| --- | --- | --- | --- | --- | --- | --- | --- | --- |
| **Demographics** |  |  |  |  |  |  |  |  |
| Age (months), n (%) |  |  |  |  |  |  |  |  |
| 6-11 | 92 (23.7) | 24 (13.7) | 25 (14.5) | 39 (16.3) | 91 (24.7) | 61 (26.9) | 53 (13.4) | 385 (19.6) |
| 12-23 | 197 (50.8) | 97 (55.4) | 88 (50.9) | 156 (65.0) | 195 (53.0) | 115 (50.7) | 234 (58.9) | 1,082 (55.0) |
| 24-35 | 99 (25.5) | 54 (30.9) | 60 (34.7) | 45 (18.8) | 82 (22.3) | 51 (22.5) | 110 (27.7) | 501 (25.5) |
| Female, n (%) | 179 (46.1) | 90 (51.4) | 78 (45.1) | 119 (49.6) | 190 (51.6) | 108 (47.6) | 192 (48.4) | 956 (48.6) |
| Mother completed secondary school or greater, n (%) | 114 (29.4) | 55 (31.4) | 24 (13.9) | 20 (8.4) | 89 (24.2) | 111 (48.9) | 8 (2.0) | 421 (21.4) |
| Wealth index, n (%)<sup>1</sup> |  |  |  |  |  |  |  |  |
| Quintile 1 (fewest assets) | 0 (0.0) | 2 (1.1) | 22 (12.7) | 3 (1.3) | 107 (29.1) | 105 (46.3) | 100 (25.2) | 339 (17.2) |
| Quintile 2 | 22 (5.7) | 36 (20.6) | 48 (27.7) | 68 (28.3) | 126 (34.2) | 87 (38.3) | 178 (44.8) | 565 (28.7) |
| Quintile 3 | 84 (21.6) | 53 (30.3) | 78 (45.1) | 114 (47.5) | 70 (19.0) | 34 (15.0) | 92 (23.2) | 525 (26.7) |
| Quintile 4 | 127 (32.7) | 62 (35.4) | 20 (11.6) | 54 (22.5) | 45 (12.2) | 1 (0.4) | 27 (6.8) | 336 (17.1) |
| Quintile 5 (most assets) | 155 (39.9) | 22 (12.6) | 5 (2.9) | 1 (0.4) | 20 (5.4) | 0 (0.0) | 0 (0.0) | 203 (10.3) |
| Received age-appropriate rotavirus vaccination, n (%)<sup>2</sup> | - | 121 (80.7) | 96 (100.0) | 191 (97.9) | 228 (85.7) | 169 (88.9) | 384 (98.2) | 1,189 (92.3) |
| Wasting, n (%)<sup>3</sup> |  |  |  |  |  |  |  |  |
| None | 321 (82.7) | 166 (94.9) | 160 (93.0) | 187 (77.9) | 230 (63.4) | 211 (95.0) | 268 (67.5) | 1,543 (78.8) |
| Moderate | 52 (13.4) | 7 (4.0) | 10 (5.8) | 43 (17.9) | 92 (25.3) | 10 (4.5) | 105 (26.4) | 319 (16.3) |
| Severe | 15 (3.9) | 2 (1.1) | 2 (1.2) | 10 (4.2) | 41 (11.3) | 1 (0.5) | 24 (6.0) | 95 (4.9) |
| Stunted, n (%)<sup>4</sup> | 112 (28.9) | 37 (21.1) | 61 (35.5) | 41 (17.1) | 152 (41.9) | 53 (23.9) | 92 (23.2) | 548 (28.0) |
| Underweight, n (%)<sup>5</sup> |  |  |  |  |  |  |  |  |
| None | 294 (75.8) | 161 (92.0) | 149 (86.1) | 184 (76.7) | 187 (51.0) | 202 (89.0) | 240 (60.5) | 1,417 (72.0) |
| Moderate | 76 (19.6) | 11 (6.3) | 19 (11.0) | 40 (16.7) | 112 (30.5) | 18 (7.9) | 119 (30.0) | 395 (20.1) |
| Severe | 18 (4.6) | 3 (1.7) | 5 (2.9) | 16 (6.7) | 68 (18.5) | 7 (3.1) | 38 (9.6) | 155 (7.9) |
| **Diarrhea episode severity indicators** |  |  |  |  |  |  |  |  |
| Diarrhea episode ≥ 6 days, n (%) | 240 (61.9) | 43 (24.6) | 31 (17.9) | 51 (21.3) | 195 (53.0) | 95 (41.9) | 49 (12.3) | 704 (35.8) |
| Number of days of diarrhea, Median (IQR) | 6 (5, 9) | 3 (2, 5) | 3 (2, 5) | 4 (3, 5) | 6 (4, 9) | 5 (3, 7) | 3 (2, 5) | 4 (3, 7) |
| Maximum number of stool in one day, Median (IQR) | 9 (7, 12) | 5 (4, 6) | 7 (5, 8) | 5 (4, 6) | 7 (6, 10) | 5 (4, 8) | 6 (5, 7) | 6 (5, 8) |
| Dysentery, n (%) | 129 (33.2) | 42 (24.0) | 29 (16.8) | 32 (13.3) | 115 (31.3) | 62 (27.3) | 140 (35.3) | 549 (27.9) |
| Dehydration, n (%)<sup>6</sup> |  |  |  |  |  |  |  |  |
| None | 353 (91.0) | 76 (43.4) | 167 (96.5) | 207 (86.3) | 303 (82.3) | 32 (14.1) | 369 (92.9) | 1,507 (76.6) |
| Some | 35 (9.0) | 95 (54.3) | 6 (3.5) | 31 (12.9) | 62 (16.8) | 193 (85.0) | 23 (5.8) | 445 (22.6) |
| Severe | 0 (0.0) | 4 (2.3) | 0 (0.0) | 2 (0.8) | 3 (0.8) | 2 (0.9) | 5 (1.3) | 16 (0.8) |
| Rehydrated at facility, n (%) | 386 (99.5) | 175 (100.0) | 173 (100.0) | 239 (99.6) | 363 (98.6) | 214 (94.3) | 394 (99.2) | 1,944 (98.8) |
| Any fever during episode, n (%) | 217 (55.9) | 146 (83.4) | 109 (63.0) | 122 (50.8) | 238 (64.7) | 176 (77.5) | 346 (87.2) | 1,354 (68.8) |
| Fever duration in days, Median (IQR) | 1 (0, 3) | 2 (1, 3) | 1 (0, 2) | 0 (0, 2) | 1 (0, 4) | 1 (0, 3) | 2 (1, 4) | 2 (0, 3) |
| Maximum axillary temperature, Median (IQR) | 36.9 (36.5, 37.3) | 37.2 (36.7, 37.9) | 36.7 (36.2, 37.1) | 36.7 (36.3, 37.2) | 36.6 (36.2, 37.0) | 36.8 (36.7, 37.5) | 37.1 (36.7, 37.8) | 36.8 (36.5, 37.4) |
| Confirmed temperature, Median (IQR) | 36.8 (36.4, 37.2) | 37.0 (36.7, 37.8) | 36.4 (36.1, 36.8) | 36.7 (36.3, 37.2) | 36.5 (36.2, 37.0) | 36.8 (36.6, 37.4) | 37.1 (36.7, 37.8) | 36.8 (36.4, 37.3) |
| Any vomiting during episode, n (%) | 133 (34.3) | 81 (46.3) | 67 (38.7) | 59 (24.6) | 130 (35.3) | 121 (53.3) | 140 (35.3) | 731 (37.1) |
| Vomiting duration in days, Median (IQR) | 0 (0, 2) | 0 (0, 2) | 0 (0, 1) | 0 (0, 0) | 0 (0, 1) | 1 (0, 1) | 0 (0, 1) | 0 (0, 1) |
| Maximum number of times vomiting in one day, Median (IQR) | 0 (0, 2) | 0 (0, 2) | 0 (0, 1) | 0 (0, 0) | 0 (0, 1) | 1 (0, 3) | 0 (0, 2) | 0 (0, 2) |
| Clark score behavioral signs, n (%) |  |  |  |  |  |  |  |  |
| None | 40 (10.3) | 51 (29.1) | 161 (93.1) | 220 (91.7) | 260 (70.7) | 60 (26.4) | 379 (95.5) | 1,171 (59.5) |
| Irritable | 347 (89.4) | 122 (69.7) | 8 (4.6) | 20 (8.3) | 105 (28.5) | 165 (72.7) | 14 (3.5) | 781 (39.7) |
| Lethargic/Unconscious | 1 (0.3) | 2 (1.1) | 0 (0.0) | 0 (0.0) | 3 (0.8) | 1 (0.4) | 4 (1.0) | 11 (0.6) |
| Convulsions | 0 (0.0) | 0 (0.0) | 4 (2.3) | 0 (0.0) | 0 (0.0) | 1 (0.4) | 0 (0.0) | 5 (0.3) |
| **Dehydration signs<sup>7</sup>** |  |  |  |  |  |  |  |  |
| Child condition, n (%) |  |  |  |  |  |  |  |  |
| Well/Alert | 40 (10.3) | 51 (29.1) | 165 (95.4) | 220 (91.7) | 260 (70.7) | 60 (26.4) | 379 (95.5) | 1,175 (59.7) |
| Restless/Irritable | 347 (89.4) | 122 (69.7) | 8 (4.6) | 20 (8.3) | 105 (28.5) | 166 (73.1) | 14 (3.5) | 782 (39.7) |
| Lethargic or Unconscious | 1 (0.3) | 2 (1.1) | 0 (0.0) | 0 (0.0) | 3 (0.8) | 1 (0.4) | 4 (1.0) | 11 (0.6) |
| Sunken eyes, n (%) | 0 (0.0) | 49 (28.0) | 6 (3.5) | 38 (15.8) | 55 (14.9) | 167 (73.6) | 32 (8.1) | 347 (17.6) |
| Ability to drink, n (%) |  |  |  |  |  |  |  |  |
| Normal/Not Thirsty | 352 (90.7) | 87 (49.7) | 168 (97.1) | 138 (57.5) | 301 (81.8) | 46 (20.3) | 375 (94.5) | 1,467 (74.5) |
| Drinks eagerly/Thirsty | 36 (9.3) | 86 (49.1) | 5 (2.9) | 101 (42.1) | 65 (17.7) | 181 (79.7) | 20 (5.0) | 494 (25.1) |
| Drinks poorly/Unable to drink | 0 (0.0) | 2 (1.1) | 0 (0.0) | 1 (0.4) | 2 (0.5) | 0 (0.0) | 2 (0.5) | 7 (0.4) |
| Skin pinch, n (%) |  |  |  |  |  |  |  |  |
| Immediately | 374 (96.4) | 144 (82.3) | 164 (94.8) | 227 (94.6) | 345 (94.0) | 108 (47.6) | 370 (93.2) | 1,732 (88.1) |
| 1 to <2 seconds (slowly) | 14 (3.6) | 29 (16.6) | 8 (4.6) | 11 (4.6) | 22 (6.0) | 118 (52.0) | 23 (5.8) | 225 (11.4) |
| 2+ seconds (very slowly) | 0 (0.0) | 2 (1.1) | 1 (0.6) | 2 (0.8) | 0 (0.0) | 1 (0.4) | 4 (1.0) | 10 (0.5) |

<sup>1</sup> Wealth index quintile calculated from country-specific wealth assessments derived from Demographic Health Surveys as detailed in Chakraborty, GHSP, 2016.
<sup>2</sup> Defined as having received all age-appropriate doses of rotavirus vaccination according to national vaccination schedules, allowing a one month buffer. Note that rotavirus is not included in the Bangladesh childhood vaccine schedule.
<sup>3</sup> Moderate wasting defined as a weight-for-height z-score of < -2 or a mid-upper arm circumference of <12.5 cm and severe wasting defined as a weight-for-height z-score of < -3 or a mid-upper arm circumference of <11.5 cm.
<sup>4</sup> Stunting defined as a height-for-age z-score of < -2.
<sup>5</sup> Moderate underweight defined as a weight-for-age z-score of < -2 and severe underweight defined as a weight-for-age z-score of < -3.
<sup>6</sup> Defined according to World Health Organization Integrated Management of Childhood Illness (WHO IMCI).
<sup>7</sup> Components of WHO IMCI dehydration definition.

Compared to all MAD, episodes of *Shigella*-attributable MAD had higher frequency of dysentery (27.9% versus 13.0%), longer duration (27.8% versus 35.8%), similar frequency of fever (68.8% versus 67.2%), and lower frequency of vomiting (37.1% versus 44.3%) and dehydration (23.4% versus 24.7%) (Figure A1). There was a higher proportion of episodes classified as moderate to severe among *Shigella*-attributable MAD compared to all MAD for all severity definitions except MVS (Table A3). Of the 1,968 children with *Shigella*-attributable MAD, 1,040 (52.8%) received guideline-recommended antibiotics.

Among *Shigella*-attributable MAD cases, the *Shigella* mortality score classified the smallest percentage of cases as moderate or severe (12.8%) while MVS +/− dysentery and MAL-ED classified the highest percentage of cases (54.3% and 53.6%, respectively). There was variation by site. Across severity definitions, children with *Shigella*-attributable MAD in Malawi and Mali had lower frequency of moderate or severe disease, whereas frequency was highest in Peru (Figure 1). A slightly higher proportion of younger children and culture-positive children (Figure A2) were classified as having moderate or severe *Shigella-*attributable MAD across severity definitions, most prominently among the definitions that include dysentery (MVS +/− dysentery and GEMS MSD).

**Figure 1.**
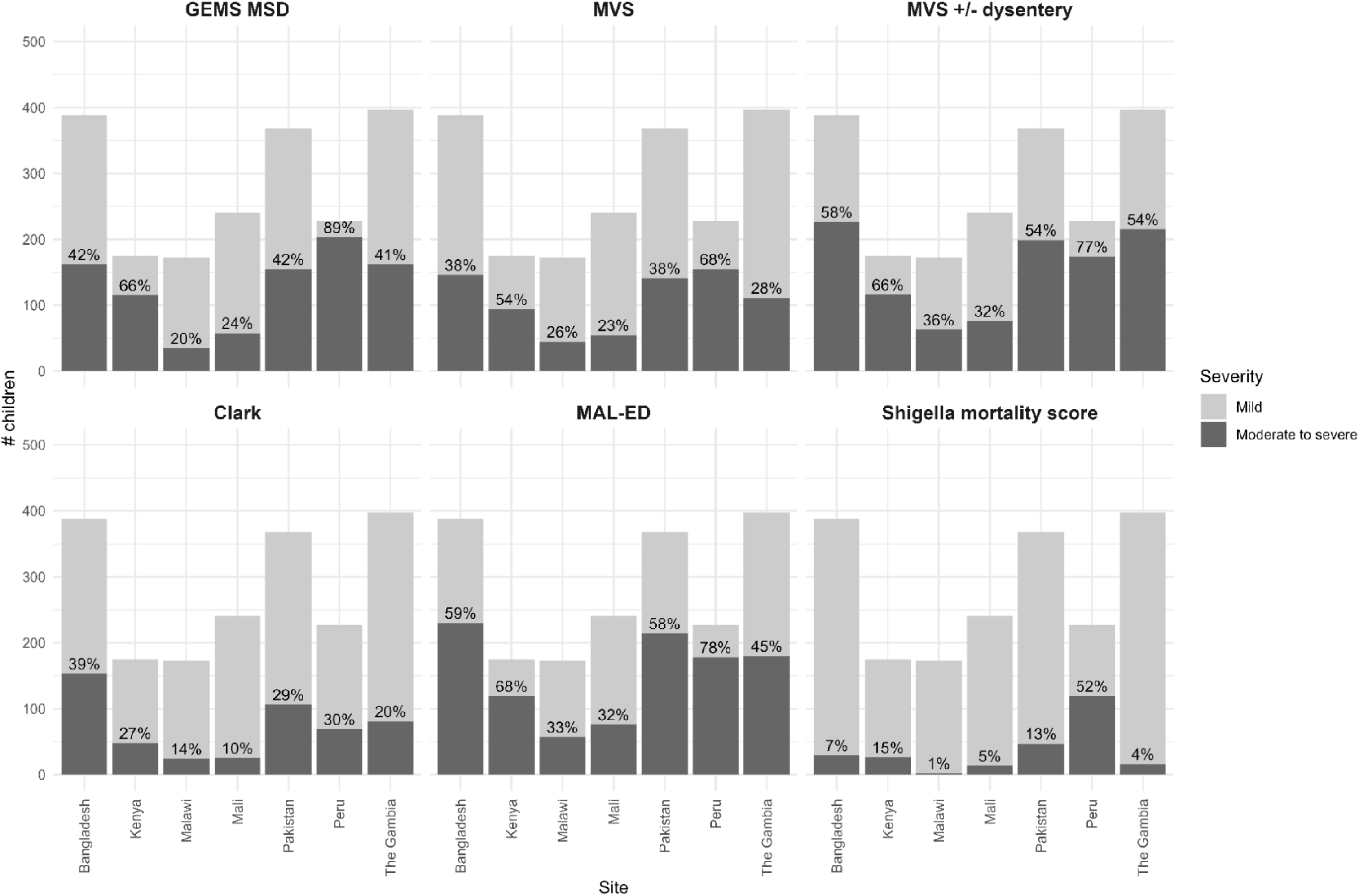
Severity of *Shigella*-attributable medically attended diarrhea by severity definition and site. Percentages indicate moderate to severe diarrhea. GEMS: Global Enteric Multicenter Study, MSD: moderate-to-severe diarrhea, MVS: modified Vesikari score Scores dichotomized as follows: MVS dichotomized as mild (<9 points) vs. moderate or severe (≥9 points); MVS +/− dysentery dichotomized as mild with no dysentery vs. moderate/severe or dysentery; GEMS MSD dichotomized as less-severe-diarrhea vs. moderate-to-severe-diarrhea; *Shigella* mortality score dichotomized as mild (<6 points) vs. moderate or severe (≥6 points) diarrhea; Clark score dichotomized as mild (<9 points) vs. moderate to severe diarrhea (≥9 points); MAL-ED dichotomized as non-severe (<6 points) vs. severe diarrhea (≥6 points) Alt text: Bar graph depicting the percentage of *Shigella*-attributable medically attended diarrhea classified as moderate or severe by each severity definition by site.

Among children with *Shigella*-attributable MAD, moderate or severe diarrhea was significantly associated with ΔLAZ/HAZ from enrollment to three-month follow-up by three severity definitions (Figure 2). Children with moderate or severe *Shigella* by MVS had an estimated ΔLAZ/HAZ that was 0.07 z-scores lower (95% CI −0.10 to −0.03) compared to children with mild MVS, while children with more severe diarrhea by MVS +/− dysentery and MAL-ED scoring also were associated with reduced ΔLAZ/HAZ of −0.05 (95% CI −0.09 to −0.02) and −0.04 (95% CI −0.07 to 0.00), respectively. For these three definitions, the magnitude of the association was larger among children with *Shigella* who did not receive guideline-recommended antibiotics, with estimated reduction in ΔLAZ/HAZ of −0.10 (95% CI −0.15 to −0.05) for severe episodes by MVS, −0.10 (95% CI −0.15 to −0.05) by MVS +/− dysentery, and −0.05 (95% CI −0.10 to 0.00) by the MAL-ED score. Similar associations were observed among all participants with MAD (Table A4).

**Figure 2.**
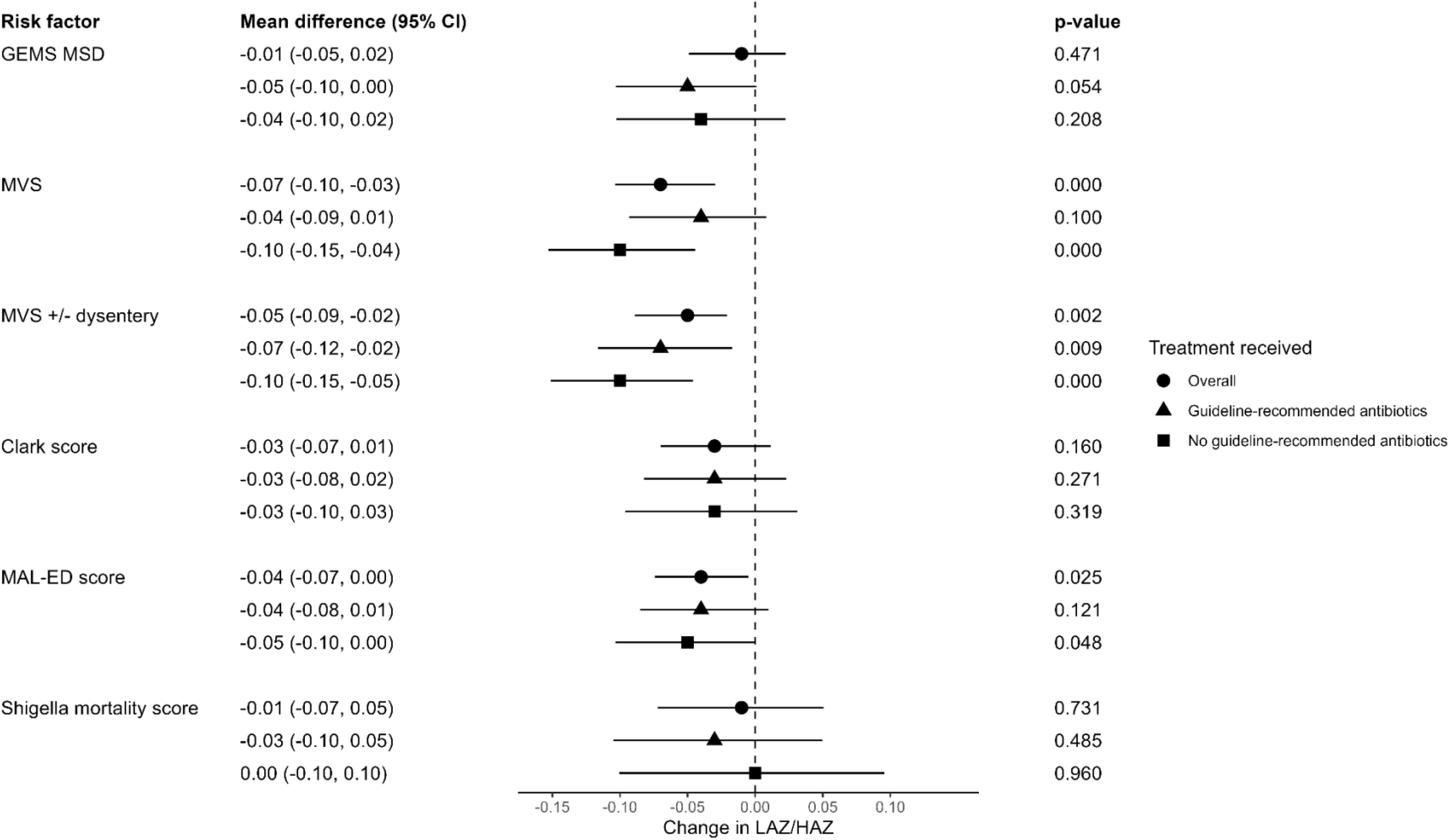
Association between more severe diarrhea by dichotomized severity definitions and change in length/ height-for-age z-score (ΔLAZ/HAZ) from enrollment to 3-month follow-up among *Shigella*-attributable medically attended diarrhea, overall and stratified by guideline-recommended antibiotic treatment. Models adjusted for baseline LAZ/HAZ, site, and number of days from enrollment to 3-month follow-up visit. GEMS: Global Enteric Multicenter Study, MSD: moderate-to-severe diarrhea, MVS: modified Vesikari score Scores dichotomized as follows: MVS dichotomized as mild (<9 points) vs. moderate or severe (≥9 points); MVS +/− dysentery dichotomized as mild with no dysentery vs. moderate/severe or dysentery; GEMS MSD dichotomized as less-severe-diarrhea vs. moderate-to-severe-diarrhea; *Shigella* mortality score dichotomized as mild (<6 points) vs. moderate or severe (≥6 points) diarrhea; Clark score dichotomized as mild (<9 points) vs. moderate to severe diarrhea (≥9 points); MAL-ED dichotomized as non-severe (<6 points) vs. severe diarrhea (≥6 points) Alt text: Forest plot depicting the association between each dichotomized severity definition and change in length/height-for-age z-score from enrollment to three-month follow-up among all *Shigella*-attributable medically attended diarrhea cases and stratified by receipt of guideline-recommended antibiotics. Among all *Shigella* cases, MVS, MVS +/− dysentery, and the MAL-ED score are negatively associated with change in length/height-for-age z-score.

Among children with *Shigella*-attributable MAD, there was one death (<0.1%) and 46 hospitalizations (2.3%) within 14 days of enrollment. The sensitivity of dichotomized severity definitions to predict hospitalization or mortality varied from 66.0% (Clark score and *Shigella* mortality score) to 93.6% (GEMS MSD) (Figure 3). The MAL-ED score (47.3%) had the lowest specificity, while the *Shigella* mortality score had the highest specificity (88.5%). When subset to children with *Shigella*-attributable MAD who did not receive guideline-recommended antibiotics, GEMS MSD and the MAL-ED score had the highest sensitivity for predicting hospitalization or mortality (90.9% and 90.9%, respectively), while the *Shigella* mortality score had the highest specificity (87.6%). Sensitivity and specificity among all MAD were similar to those among *Shigella*-attributable MAD (Table A5).

**Figure 3.**
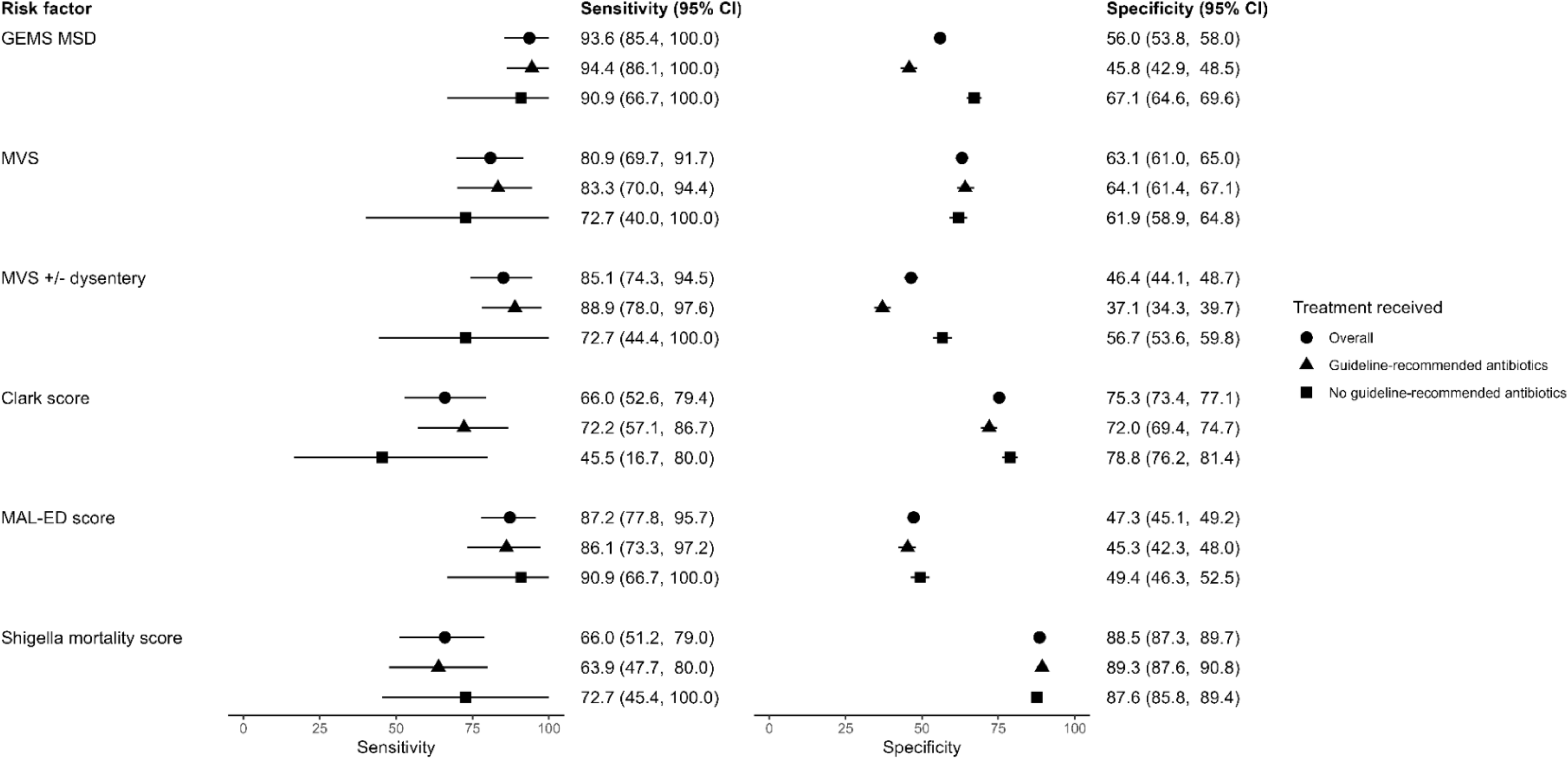
Sensitivity and specificity of dichotomized diarrhea severity scores in identifying hospitalization and/or mortality within 14 days of enrollment among *Shigella*-attributable medically-attended diarrhea. Calculated among all diarrhea cases and stratified by guideline-recommended antibiotic treatment. GEMS: Global Enteric Multicenter Study, MSD: moderate-to-severe diarrhea, MVS: modified Vesikari score Scores dichotomized as follows: MVS dichotomized as mild (<9 points) vs. moderate or severe (≥9 points); MVS +/− dysentery dichotomized as mild with no dysentery vs. moderate/severe or dysentery; GEMS MSD dichotomized as less-severe-diarrhea vs. moderate-to-severe-diarrhea; *Shigella* mortality score dichotomized as mild (<6 points) vs. moderate or severe (≥6 points) diarrhea; Clark score dichotomized as mild (<9 points) vs. moderate to severe diarrhea (≥9 points); MAL-ED dichotomized as non-severe (<6 points) vs. severe diarrhea (≥6 points) Alt text: Two side-by-site forest plots showing the sensitivity and specificity, respectively, of each dichotomized severity definition in predicting hospitalization or mortality within 14 days of enrollment. Estimates calculated among all *Shigella*-attributable medically attended diarrhea cases and stratified by receipt of guideline-recommended antibiotics. GEMS MSD, MAL-ED, MVS, and MVS +/− dysentery have the highest sensitivity, while *Shigella* mortality score and Clark score have the highest specificity.

We calculated ROC curves among *Shigella*-attributable MAD for the four numeric scores. *Shigella* mortality had the largest area under the curve ([AUC] 91.0, 95% CI 86.0 to 94.9), while MVS (79.4, 95% CI 73.4 to 85.1), Clark (76.7, 95% CI 70.3 to 83.2), and MAL-ED (75.8, 95% CI 69.7 to 82.0) had similar AUCs (Figure 4). In separate sensitivity analyses, we removed hospitalization and dehydration, from the severity definitions when present (Figure A3). Without hospitalization the *Shigella* mortality score had a smaller AUC (61.7, 95% CI 54.1 to 69.9), while MVS (73.7, 95% CI 66.8 to 80.2), Clark (76.8, 95% CI 70.3 to 83.2), and MAL-ED (75.8, 95% CI 69.7 to 82.0) had AUCs similar to the original scores. Without dehydration, each of the scores performed similarly to the original scores, with AUCs of 94.6 (95% CI 89.5 to 98.1), 79.1 (95% CI 73.4 to 84.3), 76.8 (95% CI 70.3 to 83.2), and 73.9 (95% CI 67.3 to 80.5) for *Shigella* mortality, MVS, Clark, and MAL-ED, respectively.

**Figure 4.**
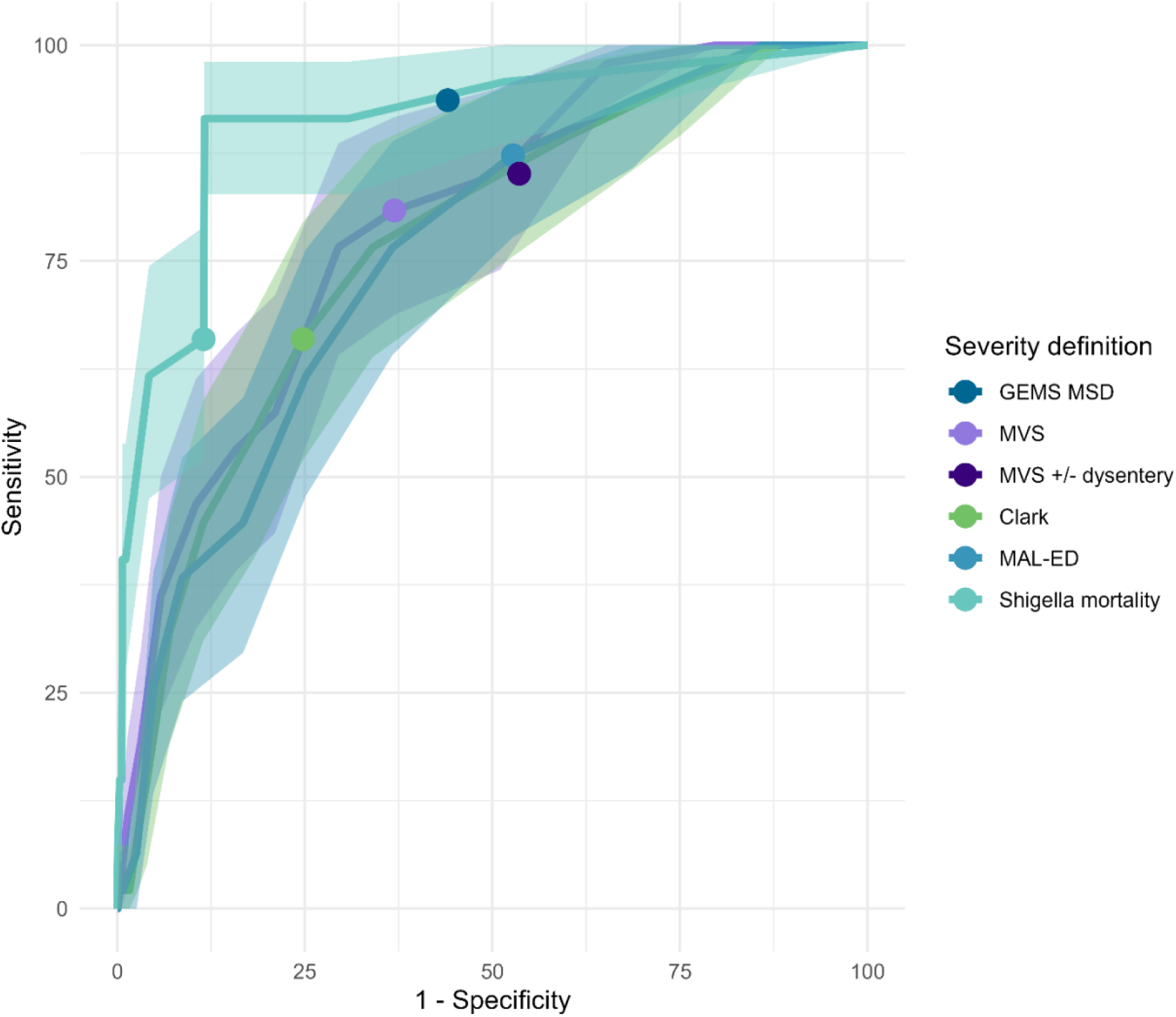
Receiver operating characteristic (ROC) curves of four continuous severity scores on the outcome of hospitalization or mortality within 14 days of enrollment among children with *Shigella*-attributable medically-attended diarrhea. Shaded bands represent 95% confidence intervals. Plotted points represent sensitivity and specificity of threshold used for dichotomizing four continuous scores, as well as sensitivity and specificity of two non-numeric severity definitions. GEMS: Global Enteric Multicenter Study, MSD: moderate-to-severe diarrhea, MVS: modified Vesikari score Scores dichotomized as follows: MVS dichotomized as mild (<9 points) vs. moderate or severe (≥9 points); MVS +/− dysentery dichotomized as mild with no dysentery vs. moderate/severe or dysentery; GEMS MSD dichotomized as less-severe-diarrhea vs. moderate-to-severe-diarrhea; *Shigella* mortality score dichotomized as mild (<6 points) vs. moderate or severe (≥6 points) diarrhea; Clark score dichotomized as mild (<9 points) vs. moderate to severe diarrhea (≥9 points); MAL-ED dichotomized as non-severe (<6 points) vs. severe diarrhea (≥6 points) Alt text: Receiver operating characteristic curves of four continuous severity scores on hospitalization or mortality within 14 days of enrollment among children with *Shigella*-attributable medically-attended diarrhea. *Shigella* mortality score has the highest area under the curve, and the other three severity scores (MVS, Clark score, and MAL-ED score) have similar curves with overlapping confidence interval bands.

## Conclusions

We evaluated the ability of six diarrhea severity definitions to disaggregate children with and without more severe *Shigella*-attributable MAD. *Shigella*-attributable MAD was more severe than overall MAD, underscoring its clinical and public health relevance and providing motivation for a *Shigella* vaccine. The proportion of *Shigella*-attributable MAD classified as moderate or severe varied across definitions, which has implications for vaccine trial sample sizes. Moderate or severe *Shigella-* attributable MAD identified using three of the six definitions—MVS, MVS +/− dysentery, and MAL-ED— was significantly associated with ΔLAZ/HAZ. Finally, most severity definitions demonstrated high sensitivity in predicting hospitalization or death, important clinical outcomes that vaccine candidates aim to prevent.

An ideal severity definition for *Shigella* vaccine trials identifies the vaccine-preventable subset of *Shigella* cases, which is often the more severe cases of disease[6,25]. The *Shigella* mortality score classified the fewest children as having moderate or severe diarrhea in our study, followed by Clark, however these two definitions did not outperform other definitions. MVS +/− dysentery and the MAL-ED score were more inclusive definitions that also performed well at predicting hospitalization, death, and linear growth loss.

Children with *Shigella*-attributable MAD had a higher proportion of moderate or severe diarrhea than all MAD by every severity definition except MVS. Fever, vomiting, dehydration, longer duration of illness, and more than six stools per 24 hours were common among children with *Shigella*-attributable MAD, all features likely to be important in classifying severe shigellosis beyond dysentery alone. Efforts to differentiate *Shigella* from other etiologies[26,27], such as viral etiologies, have found vomiting to be less common among children with *Shigella*. However, we identified vomiting in >30% of *Shigella*-attributable MAD cases, making it a viable feature to include in a severity definition among children with shigellosis. Children with culture-confirmed *Shigella* had more severe diarrhea by definitions that include dysentery compared to qPCR-attributable but culture negative cases. Culture confirmed *Shigella* cases were more likely to present with dysentery than qPCR-attributable cases[11], which may reflect the timing of disease progression identified by qPCR versus culture or the pathogen load required for detection by culture.

The considerable variation in severity classification by site was at least partially driven by differences in some or severe dehydration. Despite concerns that the components of the dehydration classification are subjective[28], no single component drove site differences in frequency of dehydration and the removal of dehydration did not substantially change the ability of severity definitions to predict hospitalization or mortality. Variation in severity by site could be due to differences in age distribution of cases, prevalent serotypes, and care-seeking behaviors[29,30]. For instance, in Peru, *Shigella* was more prevalent among the youngest age group, which tends to present with more severe disease[11]. Measuring dehydration is complex and future work may consider collecting data on alternative measures of dehydration such as the DHAKA score, which was found to better predict percent dehydration[28].

Because *Shigella* is frequently implicated in linear growth faltering, the precursor to stunting, the ideal clinical endpoint for severe shigellosis would include children at risk for *Shigella*-induced growth faltering. We found that severity as measured by MVS, MVS +/− dysentery, and the MAL-ED score was significantly associated with linear growth faltering, irrespective of antibiotic use, while severity measured by the *Shigella* mortality score, GEMS MSD, and Clark score showed non-significant associations of lesser magnitude. The three scores with significant associations included diarrhea duration as a component, a well-known risk factor for stunting[31]. This suggests that the severity definition used in a vaccine trial should at least include a component related to diarrhea duration to capture the *Shigella* infections most likely to lead to growth faltering.

We considered the composite outcome of hospitalization or mortality within 14 days of enrollment due to the small number of deaths among *Shigella*-attributable MAD cases (n=1) and because previous research has found hospitalization to be both a proxy for mortality[11] and associated with health care system costs[32]. Acknowledging there are severe cases of *Shigella* that do not lead to hospitalization or death, we focused on sensitivity when comparing definitions and found that four definitions demonstrated high sensitivity in predicting hospitalization or death: GEMS MSD, MVS, MVS +/− dysentery, and MAL-ED. To address the challenge of hospitalization being present in several severity definitions, we performed a sensitivity analysis in ROC curves removing hospitalization as a component in the four definitions where it is included. The modified versions of MVS and MVS +/− dysentery performed similarly to the original definitions, whereas the modified version of the two definitions where hospitalization was heavily weighted (GEMS MSD and *Shigella* mortality score) did not perform as well. MVS, MVS +/− dysentery, and the MAL-ED score were the most sensitive scores in predicting hospitalization and mortality and the most robust to the removal of hospitalization. Previous research found that MVS and the MAL-ED score were associated with hospitalization among children enrolled at an emergency department[13] and in the community[15], respectively, and our study now builds upon these findings to suggest that these scores may be useful proxies for hospitalization or death among *Shigella* cases in LMICs.

This large, multi-site study utilized detailed clinical data to compare diarrhea severity definitions among children with *Shigella* across diverse settings. Given the three-month follow-up period, we examined hospitalization, mortality, and growth faltering as these are the most serious measured outcomes that a vaccine may avert but acknowledge that the shorter follow-up time may have limited our ability to fully assess growth faltering. In addition, the overlap between severity definitions and the outcome of hospitalization or mortality limited our ability to comprehensively evaluate the two definitions where hospitalization was weighted heavily, GEMS MSD and *Shigella* mortality score. The magnitude of linear growth faltering observed in this study may not be clinically meaningful on an individual level but is similar in magnitude to highly referenced findings in the *Shigella* growth literature[24]. Finally, antibiotic treatment is likely to be contributing to the relationship between severity and clinical outcomes, although our analyses stratifying by appropriate antibiotic treatment attempted to overcome this.

We saw similar performance across three severity definitions (MVS, MVS +/− dysentery, and the MAL-ED score) examining two types of adverse outcomes from *Shigella*. Of these definitions, only MVS +/− dysentery contains dysentery, a clear marker of *Shigella* clinical severity that should be included in a vaccine trial case definition. Diarrhea duration has been previously linked with mortality[11] and may improve prediction of changes in LAZ/HAZ, indicating it should be considered in the severity definition used for eventual *Shigella* vaccine trials. Other practical considerations in severity definition selection include the trade-offs between simpler definitions, which are easy to understand and may only require data collection at one point in time, versus scores, which can be further disaggregated but may require follow-up data collection[14]. There are multiple viable existing options for a moderate or severe *Shigella* case definition that balance association with relevant adverse outcomes with requisite underlying frequency in populations of interest.

## Data Availability

The EFGH statistical analysis plan (https://clinicaltrials.gov/study/NCT06047821) and study protocol (https://academic.oup.com/ofid/issue/11/Supplement_1) were made publicly available. The datasets were deidentified and anonymized and will be publicly available upon publication of the manuscript.

## Acknowledgements

The authors thank the children who participated in these studies and their families, and the dedicated physicians, nurses, scientists, and staff at each study site for their dedication and outstanding performance of clinical and laboratory study activities.

## Methods appendix

### Key definitions

#### Diarrhea episode

The diarrhea episode was defined as the period from diarrhea start to the last day of diarrhea followed by two diarrhea-free days. The first day of the index diarrhea episode was identified by caregivers during the screening process. The last day of the episode was determined using the diarrhea diary and follow-up visits, when caregivers were asked about the last day of the participant’s episode. The number of loose stools was recorded daily in the diarrhea diary, and the last day of the episode was determined by the last day with at least three recorded loose stools, followed by two days of less than three loose stools. When the diarrhea episode was still ongoing at the end of the diarrhea diary (14 days after enrollment), the last date of the diarrhea episode reported at the follow-up visit was used. If the last date reported at follow-up was missing or prior to 14 days after enrollment, then the episode was assumed to have ended on the last day of the diarrhea diary. In cases where the diarrhea diary was incomplete, any days with missing values for loose stool count were assumed to have less than three loose stools. When no diarrhea diary was submitted, the diarrhea end date reported by caregivers at the follow-up visit was used. If both the diarrhea diary and follow-up date were missing, the diarrhea episode was assumed to end on the day of enrollment.

#### Number of days of diarrhea

The number of days of diarrhea was defined as the number of days within the diarrhea episode where the caregiver reported three or more loose stools.

#### Diarrhea duration prior to presentation

The number of days from diarrhea episode start to enrollment date (including the day of enrollment).

#### Maximum number of loose stools per day

At enrollment and follow-up visits, caregivers are asked to report the maximum number of loose stools their child has passed in a single day during the index episode. The number of loose stools per day from enrollment to the end of the episode is also recorded in the diarrhea diary. The maximum number of loose stools in a 24-hour period is taken from the maximum of these three sources.

#### Dysentery

Caregivers were asked to report if they observed blood in the participant’s stool at any point during the index diarrhea episode at screening, enrollment, and follow-up visits. Caregivers also recorded the presence of blood in stool daily for 14 days following enrollment in the diarrhea diary. Additionally, clinician diagnosis of dysentery was recorded at enrollment, both at presentation and discharge from the health facility. If responses from caregivers or clinician diagnosis at any time point indicated that the child had blood in stool during the index episode, the child was classified as having dysentery.

#### Dehydration

All participants were classified as having none, some, or severe dehydration at enrollment using World Health Organization (WHO) dehydration categories from the Integrated Management of Childhood Illness guidelines[33]. A child with two or more of the following symptoms was classified as having severe dehydration: lethargic or unconscious, abnormally sunken eyes observed by clinician and confirmed by caregiver, child drinks poorly or is unable to drink, and skin pinch returns to normal after 2 or more seconds. A child who does not meet the criteria for severely dehydrated and has two or more of the following symptoms was classified as having some dehydration: restless or irritable, abnormally sunken eyes observed by clinician and confirmed by caregiver, child drinks eagerly or is thirsty, and skin pinch returns to normal in 1 to 2 seconds. A child who does not meet the criteria for either some or severe dehydration is classified as having none.

#### Rehydration treatment

A participant was considered to have received rehydration treatment at enrollment if they received oral rehydration solution or IV rehydration at the health facility or if they were instructed to continue oral rehydration treatment at home upon discharge.

#### Maximum axillary or rectal temperature

Temperature was measured by clinicians at enrollment. Caregivers with a thermometer at home were instructed to take and record their child’s temperature in the diarrhea diary daily from enrollment through 14 days after enrollment. At the follow-up visit, caregivers were asked to report the highest temperature recorded during the diarrhea episode if the child had been feverish. Temperatures taken via armpit, forehead, or unknown location were classified as axillary temperatures. Oral temperature was converted to axillary by subtracting 0.5°C, and rectal temperature was converted to axillary temperature using the following formula: axillary temperature = 0.94 * rectal temperature + 2.92. The maximum axillary temperature was calculated for each child by taking the highest recorded temperature from any source during the diarrhea episode. This was used to calculate the maximum rectal temperature using the above formula.

#### Confirmed temperature

Confirmed temperature was the temperature taken at enrollment by a clinician.

#### Duration of fever

At enrollment, caregivers are asked how many days the child has felt hot to the touch since the beginning of the diarrhea episode. Following enrollment, temperature data is recorded via the diarrhea diary and follow-up visits. For each day in the diarrhea diary, the child is considered feverish if they have an oral temperature of ≥37.8°C or an axillary temperature of ≥37.2°C or, if temperature is not measured, if the caregiver records that the child feels “hot to touch”. If no temperature is taken and the caregiver does not record a “hot to touch” indication, the child is assumed to not have fever for that day. Separately from the diarrhea diary, caregivers are asked during follow-up visits how many days the child had a fever from enrollment until the end of the index diarrhea episode. The number of days a participant has fever from enrollment until the end of the episode is taken from the diarrhea diary except when the diarrhea diary is missing or when the follow-up response indicates more feverish days than the diarrhea diary. In those cases, the number of days indicated by caregiver’s follow-up response is used. The number of days of fever or hot to touch assessment for the entire diarrhea episode is calculated by adding the number of days reported at enrollment to the number of days calculated by either the diary or follow-up visits.

#### Maximum number of times vomiting in a day

Caregivers reported the maximum number of times the child vomited in a day since the beginning of the diarrhea at enrollment. Caregivers then recorded the number of times their child vomited daily from enrollment through 14 days after enrollment in the diarrhea diary. At the follow-up visit, caregivers were asked to report the maximum number of times the child vomited in a day from enrollment until the end of the episode. If the diarrhea diary was returned, the maximum number of times vomited in a day was the highest recorded value from the enrollment and diarrhea diary during the diarrhea episode. If no diarrhea diary was returned, then the maximum number of times vomited in a day was the highest recorded value from the enrollment and follow-up responses.

#### Duration vomiting

At enrollment, caregivers are asked how many days the child has vomited since the beginning of the diarrhea episode. The number of days reported at enrollment is summed with the number of days the child has vomiting during the episode as indicated in the diarrhea diary. When data in the diary is missing, it is assumed no vomiting occurred that day.

#### Behavioral signs

Behavioral signs are categorized based on clinician assessment at enrollment. A clinician report of a child experiencing convulsions were classified as having “Seizures”; here, we used convulsions as a proxy for seizures. Children who were not experiencing convulsions and were reported to be lethargic or unconscious were classified as “Lethargic/Listless”, and those who were not experiencing convulsions and were reported to be restless or irritable were classified as “Irritable/Less playful”.

#### Guideline-recommended antibiotics

Children who received azithromycin, ciprofloxacin, ceftriaxone, or pivmecillinam during the diarrhea episode were classified as having received guideline-recommended antibiotics, based on WHO recommendations for *Shigella* treatment[23]. All other children were classified as having not received guideline-recommended antibiotics.

#### Hospitalization

was assessed at enrollment and follow-up visits by caregiver recall and hospital records (gold standard) when available. Date and time of admission, length of hospital stay, presenting signs/symptoms, and treatment received were obtained when available. Hospitalization was defined as an overnight stay (child was on the ward from at least 12am to 6am or self-discharged prior to that timeframe). Hospitalizations missing admission date (n=7) were excluded from analysis.

#### Deaths

occurring within the three-month follow-up period were assessed by caregiver report at each scheduled visit or during upcoming visit phone reminders. Date and cause of death were obtained from caregiver history, hospital records or death certificate, when available. The death certificate was considered the gold standard for date of death.

#### Length/Height-for-age z-score (LAZ/HAZ)

The LAZ/HAZ was calculated for each child at enrollment, 4-week follow-up visit, and 3-month follow-up visit according to WHO guidelines[20]. Length (<24 months of age) or height (≥24 months of age) was measured during each visit to the nearest 0.1 cm[16]. The average of two measurements was used, and if the first two measurements had a difference of >0.5 cm a third measurement was taken. Each study team received initial training and conducted anthropometry standardization tests every 6 months, which guided subsequent refresher trainings. At least 5% of children at each site were re-measured by a gold standard measurer for quality control.

### Severity definitions

#### Modified Vesikari Score (MVS)[12]

The MVS is a summed score with the following components: number of days of diarrhea of 1-4 days is 1 point, 5 days is 2 points, and ≥6 days is 3 points; a maximum of 1-3 loose stools in a 24-hour period is 1 point, 4-5 loose stools is 2 points, and ≥6 loose stools is 3 points; duration of vomiting of 1 day is 1 point, 2 days is 2 points, and ≥3 days is 3 points; a maximum of 1 vomiting episode in a 24-hour period is 1 point, 2-4 episodes is 2 points, and ≥5 episodes is 3 points; a maximum axillary temperature of 36.6-37.9°C is 1 point, 38.0-38.4°C is 2 points, and ≥38.5°C is 3 points; some dehydration at enrollment is 2 points and severe dehydration at enrollment is 3 points; and rehydration treatment is 1 point and hospitalization is 2 points. The scores will be summed and categorized as mild illness (0-8 points), moderate illness (9-10 points), and severe illness (11+ points).

#### Moderate to severe MVS +/− dysentery[14]

Moderate or severe diarrhea by MVS or dysenterywill be defined as a MVS of 9 or more points or presence of dysentery, defined as above. Participants with an MVS score of 8 or less who do not have dysentery will be classified as mild with no dysentery.

#### GEMS moderate-to-severe diarrhea (MSD)[1]

GEMS MSD is defined as presenting to a health facility with diarrhea and at least one of the following: some or severe dehydration by WHO IMCI criteria, dysentery, or inpatient admission. Less-severe-diarrhea (LSD) is defined as presenting to a health facility for diarrhea without MSD.

#### *Shigella* mortality score[11]

*Shigella* mortality score is a summed score with the following components: duration of diarrhea episode of 1-3 days is 0 points, 4-5 days is 2 points, and ≥6 days is 3 points; WHO-defined dehydration classification of some dehydration at enrollment is 4 points, and severe dehydration at enrollment is 8 points; and inpatient admission is 5 points. The scores will be summed and categorized as mild (<6 points), moderate (6-8 points), and severe (9+ points).

#### Clark score[22]

Clark score is a summed score with the following components: number of days of diarrhea of 1-4 days is 1 point, 5-7 days is 2 points, and >7 days is 3 points; a maximum of 2-4 loose stools in a 24-hour period is 1 point, 5-7 loose stools is 2 points, and >7 loose stools is 3 points; duration of vomiting of 2 days is 1 point, 3-5 days is 2 points, and >5 days is 3 points; a maximum of 1-3 vomiting episodes in a 24-hour period is 1 point, 4-6 episodes is 2 points, and >6 is 3 points; duration of reported fever of 1-2 days is 1 point, 3-4 days is 2 points, and ≥5 days is 3 points; a maximum rectal temperature of 38-38.2°C is 1 point, 38.3-38.7°C is 2 points, and ≥38.8°C is 3 points; clinician behavioral classification of irritable/less playful is 1 point, lethargic/listless is 2 points, and seizures is 3 points. The scores will be summed and categorized as mild (2-8 points) and moderate to severe (9+ points).

#### MAL-ED score[15]

MAL-ED score is a summed score with the following components: number of days of diarrhea of 2-4 days is 1 point, 5-7 days is 2 points, and ≥8 days is 3 points; a maximum of <5 loose stools in a 24-hour period is 1 point, 5-7 loose stools is 2 points, and ≥8 stools is 3 points; duration of vomiting of 1 day is 1 point, 2 days is 2 points, and ≥3 days is 3 points; duration of reported fever of ≥1 days is 1 point; clinician confirmed temperature of ≥37.5°C is 2 points; WHO-defined dehydration classification of some dehydration at enrollment is 2 points, and severe dehydration at enrollment is 3 points. The scores will be summed and categorized as non-severe (<6 points) or severe (6+ points)

### Reformulating severity definitions for sensitivity analysis

To further explore the relationship between severity definitions and the combined outcome of hospitalization or death, we performed two sensitivity analysis removing hospitalization and dehydration, respectively, from definitions which included either as a component of the definition and then rescaling scores. Four definitions include hospitalization (MVS, MVS +/− dysentery, GEMS-MSD, and the *Shigella* mortality score) and five include dehydration (MVS, MVS +/− dysentery, GEMS-MSD, the *Shigella* mortality score, and the MAL-ED score). For GEMS-MSD, hospitalization or dehydration were dropped as a criterion for moderate-to-severe diarrhea. For the four numeric scores, hospitalization or dehydration was removed from the score calculations, and a new threshold for moderate to severe disease was calculated as the same proportion of points from the original threshold to the original highest possible severity score. For MVS and MVS +/− dysentery, the treatment and dehydration components were removed, reducing the highest possible score from 20 to 18 and 17 and the threshold for moderate or severe disease from 9 to 8 and 8 points, respectively. For the *Shigella* mortality score, removing hospitalization reduced the highest possible score from 16 to 11 and the threshold for moderate or severe disease from 6 to 4 points, while removing dehydration reduced the highest possible score to 8 and the threshold for moderate or severe disease to 3 points. For the MAL-ED score, removing dehydration reduced the highest possible score from 15 to 12 and changed the threshold for moderate or severe disease from 6 to 5 points.

## Results appendix

**Table A1:**
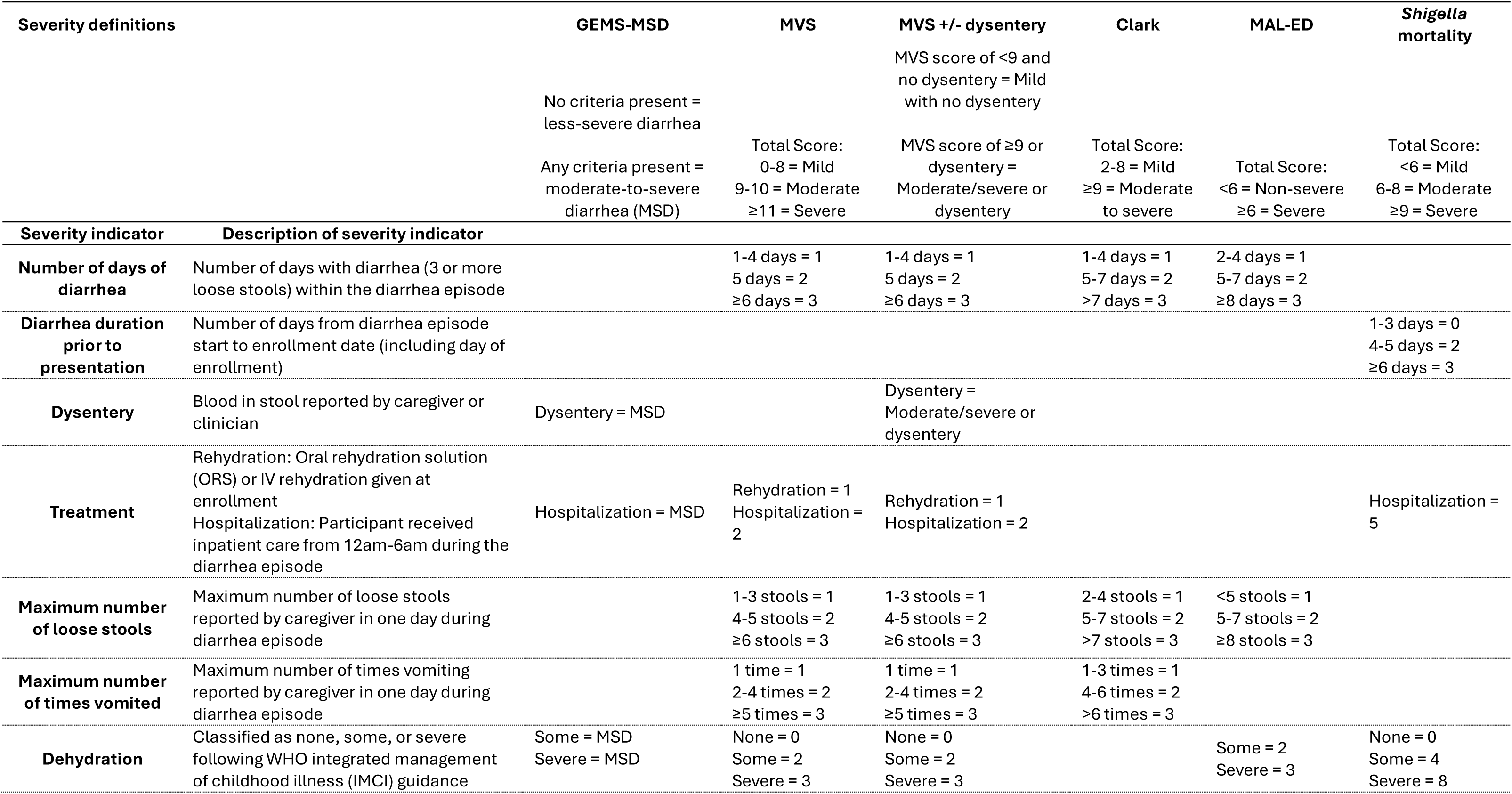
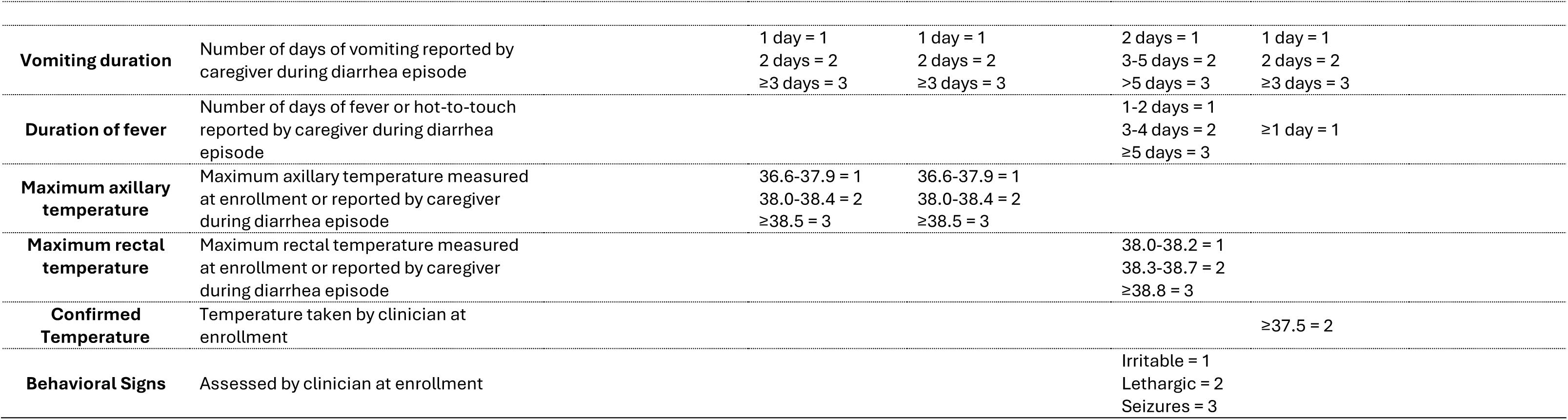
Construction of severity definitions.

**Table A2.** Demographic characteristics and diarrhea severity indicators of participants with medically-attended diarrhea (all participants) by site.

| Characteristic | Bangladesh<br>N = 1,361 | Kenya<br>N = 1,400 | Malawi<br>N = 1,399 | Mali<br>N = 1,400 | Pakistan<br>N = 1,400 | Peru<br>N = 1,117 | The Gambia<br>N = 1,399 | Overall<br>N = 9,476 |
| --- | --- | --- | --- | --- | --- | --- | --- | --- |
| **Demographics** |  |  |  |  |  |  |  |  |
| Age (months), n (%) |  |  |  |  |  |  |  |  |
| 6-11 | 572 (42.0) | 560 (40.0) | 478 (34.2) | 600 (42.9) | 454 (32.4) | 365 (32.7) | 446 (31.9) | 3,475 (36.7) |
| 12-23 | 588 (43.2) | 595 (42.5) | 627 (44.8) | 613 (43.8) | 646 (46.1) | 585 (52.4) | 718 (51.3) | 4,372 (46.1) |
| 24-35 | 201 (14.8) | 245 (17.5) | 294 (21.0) | 187 (13.4) | 300 (21.4) | 167 (15.0) | 235 (16.8) | 1,629 (17.2) |
| Female, n (%) | 596 (43.8) | 637 (45.5) | 651 (46.5) | 643 (45.9) | 655 (46.8) | 488 (43.7) | 646 (46.2) | 4,316 (45.5) |
| Mother completed secondary school or greater, n (%) | 400 (29.4) | 404 (28.9) | 221 (15.8) | 125 (9.0) | 286 (20.4) | 608 (54.4) | 58 (4.2) | 2,102 (22.2) |
| Wealth index, n (%)<sup>1</sup> |  |  |  |  |  |  |  |  |
| Quintile 1 (fewest assets) | 0 (0.0) | 34 (2.4) | 158 (11.3) | 23 (1.6) | 458 (32.7) | 507 (45.4) | 346 (24.7) | 1,526 (16.1) |
| Quintile 2 | 83 (6.1) | 247 (17.6) | 449 (32.1) | 430 (30.7) | 475 (33.9) | 430 (38.5) | 596 (42.6) | 2,710 (28.6) |
| Quintile 3 | 278 (20.4) | 404 (28.9) | 538 (38.5) | 600 (42.9) | 235 (16.8) | 173 (15.5) | 355 (25.4) | 2,583 (27.3) |
| Quintile 4 | 430 (31.6) | 522 (37.3) | 196 (14.0) | 335 (23.9) | 170 (12.1) | 7 (0.6) | 102 (7.3) | 1,762 (18.6) |
| Quintile 5 (most assets) | 570 (41.9) | 193 (13.8) | 58 (4.1) | 12 (0.9) | 62 (4.4) | 0 (0.0) | 0 (0.0) | 895 (9.4) |
| Received age-appropriate rotavirus vaccination, n (%)<sup>2</sup> | - | 814 (67.2) | 919 (96.3) | 1,168 (95.0) | 845 (85.3) | 858 (90.7) | 1,364 (98.3) | 5,968 (88.8) |
| Wasting, n (%)<sup>3</sup> |  |  |  |  |  |  |  |  |
| None | 1,119 (82.3) | 1,337 (95.7) | 1,327 (95.4) | 1,086 (77.6) | 913 (65.8) | 1,052 (95.5) | 1,000 (71.7) | 7,834 (83.1) |
| Moderate | 206 (15.1) | 48 (3.4) | 53 (3.8) | 261 (18.6) | 334 (24.1) | 44 (4.0) | 307 (22.0) | 1,253 (13.3) |
| Severe | 35 (2.6) | 12 (0.9) | 11 (0.8) | 53 (3.8) | 141 (10.2) | 6 (0.5) | 87 (6.2) | 345 (3.7) |
| Stunted, n (%)<sup>4</sup> | 331 (24.3) | 257 (18.4) | 402 (28.9) | 183 (13.1) | 540 (39.0) | 233 (21.2) | 288 (20.7) | 2,234 (23.7) |
| Underweight, n (%)<sup>5</sup> |  |  |  |  |  |  |  |  |
| None | 1,081 (79.4) | 1,297 (92.6) | 1,224 (87.5) | 1,088 (77.7) | 799 (57.2) | 1,008 (90.2) | 919 (65.7) | 7,416 (78.3) |
| Moderate | 222 (16.3) | 82 (5.9) | 139 (9.9) | 239 (17.1) | 370 (26.5) | 90 (8.1) | 351 (25.1) | 1,493 (15.8) |
| Severe | 58 (4.3) | 21 (1.5) | 36 (2.6) | 73 (5.2) | 227 (16.3) | 19 (1.7) | 129 (9.2) | 563 (5.9) |
| **Diarrhea episode severity indicators** |  |  |  |  |  |  |  |  |
| Diarrhea episode ≥ 6 days, n (%) | 779 (57.2) | 267 (19.1) | 250 (17.9) | 200 (14.3) | 569 (40.6) | 416 (37.2) | 155 (11.1) | 2,636 (27.8) |
| Number of days of diarrhea, Median (IQR) | 6 (4, 8) | 3 (2, 5) | 3 (3, 5) | 4 (3, 5) | 5 (3, 7) | 5 (3, 7) | 3 (2, 4) | 4 (3, 6) |
| Maximum number of stool in one day, Median (IQR) | 10 (7, 12) | 5 (4, 6) | 6 (5, 8) | 5 (4, 6) | 7 (6, 9) | 5 (4, 7) | 5 (4, 7) | 6 (4, 8) |
| Dysentery, n (%) | 206 (15.1) | 148 (10.6) | 138 (9.9) | 93 (6.6) | 234 (16.7) | 170 (15.2) | 241 (17.2) | 1,230 (13.0) |
| Dehydration, n (%)<sup>6</sup> |  |  |  |  |  |  |  |  |
| None | 1,191 (87.5) | 656 (46.9) | 1,348 (96.4) | 1,236 (88.3) | 1,185 (84.6) | 214 (19.2) | 1,310 (93.6) | 7,140 (75.3) |
| Some | 168 (12.3) | 685 (48.9) | 49 (3.5) | 154 (11.0) | 207 (14.8) | 897 (80.3) | 70 (5.0) | 2,230 (23.5) |
| Severe | 2 (0.1) | 59 (4.2) | 2 (0.1) | 10 (0.7) | 8 (0.6) | 6 (0.5) | 19 (1.4) | 106 (1.1) |
| Rehydrated at facility, n (%) | 1,359 (99.9) | 1,397 (99.8) | 1,399 (100.0) | 1,395 (99.6) | 1,376 (98.3) | 1,016 (91.0) | 1,390 (99.4) | 9,332 (98.5) |
| Any fever during episode, n (%) | 776 (57.0) | 1,167 (83.4) | 880 (62.9) | 720 (51.4) | 849 (60.6) | 743 (66.5) | 1,231 (88.0) | 6,366 (67.2) |
| Fever duration in days, Median (IQR) | 1 (0, 3) | 2 (1, 3) | 1 (0, 2) | 0 (0, 2) | 1 (0, 3) | 1 (0, 2) | 2 (2, 4) | 1 (0, 3) |
| Maximum axillary temperature, Median (IQR) | 36.8 (36.5, 37.2) | 37.2 (36.7, 37.9) | 36.7 (36.3, 37.2) | 36.7 (36.4, 37.2) | 36.6 (36.2, 37.0) | 36.8 (36.6, 37.3) | 37.1 (36.7, 37.7) | 36.8 (36.5, 37.4) |
| Confirmed temperature, Median (IQR) | 36.7 (36.4, 37.1) | 37.1 (36.7, 37.8) | 36.4 (36.1, 36.8) | 36.7 (36.4, 37.2) | 36.6 (36.2, 37.0) | 36.8 (36.6, 37.2) | 37.1 (36.7, 37.7) | 36.8 (36.4, 37.3) |
| Any vomiting during episode, n (%) | 648 (47.6) | 749 (53.5) | 637 (45.5) | 417 (29.8) | 562 (40.1) | 541 (48.4) | 641 (45.8) | 4,195 (44.3) |
| Vomiting duration in days, Median (IQR) | 0 (0, 2) | 1 (0, 2) | 0 (0, 2) | 0 (0, 1) | 0 (0, 1) | 0 (0, 1) | 0 (0, 2) | 0 (0, 2) |
| Maximum number of times vomiting in one day, Median (IQR) | 0 (0, 3) | 1 (0, 2) | 0 (0, 2) | 0 (0, 2) | 0 (0, 2) | 0 (0, 3) | 0 (0, 3) | 0 (0, 2) |
| Clark score behavioral signs, n (%) |  |  |  |  |  |  |  |  |
| None | 137 (10.1) | 374 (26.7) | 1,344 (96.1) | 1,293 (92.4) | 1,056 (75.4) | 354 (31.7) | 1,348 (96.4) | 5,906 (62.3) |
| Irritable | 1,221 (89.7) | 999 (71.4) | 30 (2.1) | 105 (7.5) | 331 (23.6) | 755 (67.6) | 35 (2.5) | 3,476 (36.7) |
| Lethargic/Unconscious | 3 (0.2) | 20 (1.4) | 1 (0.1) | 1 (0.1) | 13 (0.9) | 5 (0.4) | 14 (1.0) | 57 (0.6) |
| Convulsions | 0 (0.0) | 7 (0.5) | 24 (1.7) | 1 (0.1) | 0 (0.0) | 3 (0.3) | 2 (0.1) | 37 (0.4) |
| **Dehydration signs<sup>7</sup>** |  |  |  |  |  |  |  |  |
| Child condition, n (%) |  |  |  |  |  |  |  |  |
| Well/Alert | 137 (10.1) | 375 (26.8) | 1,367 (97.7) | 1,294 (92.4) | 1,056 (75.4) | 354 (31.7) | 1,350 (96.5) | 5,933 (62.6) |
| Restless/Irritable | 1,221 (89.7) | 1,005 (71.8) | 31 (2.2) | 105 (7.5) | 331 (23.6) | 758 (67.9) | 35 (2.5) | 3,486 (36.8) |
| Lethargic or Unconscious | 3 (0.2) | 20 (1.4) | 1 (0.1) | 1 (0.1) | 13 (0.9) | 5 (0.4) | 14 (1.0) | 57 (0.6) |
| Sunken eyes, n (%) | 1 (0.1) | 364 (26.0) | 58 (4.1) | 239 (17.1) | 176 (12.6) | 757 (67.8) | 96 (6.9) | 1,691 (17.8) |
| Ability to drink, n (%) |  |  |  |  |  |  |  |  |
| Normal/Not Thirsty | 1,195 (87.8) | 750 (53.6) | 1,336 (95.5) | 842 (60.1) | 1,134 (81.0) | 248 (22.2) | 1,327 (94.9) | 6,832 (72.1) |
| Drinks eagerly/Thirsty | 163 (12.0) | 612 (43.7) | 57 (4.1) | 548 (39.1) | 260 (18.6) | 868 (77.7) | 66 (4.7) | 2,574 (27.2) |
| Drinks poorly/Unable to drink | 3 (0.2) | 38 (2.7) | 6 (0.4) | 10 (0.7) | 6 (0.4) | 1 (0.1) | 6 (0.4) | 70 (0.7) |
| Skin pinch, n (%) |  |  |  |  |  |  |  |  |
| Immediately | 1,279 (94.0) | 1,196 (85.4) | 1,357 (97.0) | 1,329 (94.9) | 1,336 (95.5) | 623 (55.8) | 1,317 (94.1) | 8,437 (89.0) |
| 1 to <2 seconds (slowly) | 79 (5.8) | 181 (12.9) | 40 (2.9) | 65 (4.6) | 63 (4.5) | 492 (44.0) | 68 (4.9) | 988 (10.4) |
| 2+ seconds (very slowly) | 3 (0.2) | 23 (1.6) | 2 (0.1) | 6 (0.4) | 0 (0.0) | 2 (0.2) | 14 (1.0) | 50 (0.5) |

<sup>1</sup> Wealth index quintile calculated from country-specific wealth assessments derived from Demographic Health Surveys as detailed in Chakraborty, GHSP, 2016.
<sup>2</sup> Defined as having received all age-appropriate doses of rotavirus vaccination according to national vaccination schedules, allowing a one month buffer. Note that rotavirus is not included in the Bangladesh childhood vaccine schedule.
<sup>3</sup> Moderate wasting defined as a weight-for-height z-score of < -2 or a mid-upper arm circumference of <12.5 cm and severe wasting defined as a weight-for-height z-score of < -3 or a mid-upper arm circumference of <11.5 cm.
<sup>4</sup> Stunting defined as a height-for-age z-score of < -2.
<sup>5</sup> Moderate underweight defined as a weight-for-age z-score of < -2 and severe underweight defined as a weight-for-age z-score of < -3.
<sup>6</sup> Defined according to World Health Organization Integrated Management of Childhood Illness (WHO IMCI).
<sup>7</sup> Components of WHO IMCI dehydration definition.

**Table A3.** Proportion of children with medically-attended diarrhea (MAD) and *Shigella*-attributable MAD by severity score categories by site.

| Severity definition | Bangladesh | Kenya | Malawi | Mali | Pakistan | Peru | The Gambia | Overall |
| --- | --- | --- | --- | --- | --- | --- | --- | --- |
| **All MAD** |  |  |  |  |  |  |  |  |
| GEMS MSD, n (%)<sup>1</sup> |  |  |  |  |  |  |  |  |
| Less-severe | 966 (71.0) | 584 (41.7) | 1,214 (86.8) | 1,161 (82.9) | 989 (70.6) | 184 (16.5) | 1,062 (75.9) | 6,160 (65.0) |
| Moderate-to-severe | 395 (29.0) | 816 (58.3) | 185 (13.2) | 239 (17.1) | 411 (29.4) | 933 (83.5) | 337 (24.1) | 3,316 (35.0) |
| Modified Vesikari score (MVS), Median (IQR), n (%)<sup>2</sup> | 8 (7, 12) | 9 (6, 11) | 7 (5, 9) | 6 (5, 9) | 8 (6, 10) | 9 (7, 11) | 7 (6, 10) | 8 (6, 10) |
| Mild | 692 (50.8) | 668 (47.7) | 946 (67.6) | 1,046 (74.7) | 883 (63.1) | 456 (40.8) | 888 (63.5) | 5,579 (58.9) |
| Moderate | 175 (12.9) | 289 (20.6) | 235 (16.8) | 173 (12.4) | 239 (17.1) | 294 (26.3) | 248 (17.7) | 1,653 (17.4) |
| Severe | 494 (36.3) | 443 (31.6) | 218 (15.6) | 181 (12.9) | 278 (19.9) | 367 (32.9) | 263 (18.8) | 2,244 (23.7) |
| Moderate or severe diarrhea by MVS or dysentery, n (%)<sup>3</sup> | 796 (58.5) | 802 (57.3) | 538 (38.5) | 407 (29.1) | 644 (46.0) | 722 (64.6) | 673 (48.1) | 4,582 (48.4) |
| Clark score, Median (IQR), n (%)<sup>4</sup> | 8 (6, 11) | 7 (5, 9) | 5 (4, 7) | 4 (3, 6) | 6 (4, 9) | 6 (4, 8) | 6 (5, 8) | 6 (4, 8) |
| Mild | 770 (56.6) | 971 (69.4) | 1,176 (84.1) | 1,264 (90.3) | 1,033 (73.8) | 853 (76.4) | 1,077 (77.0) | 7,144 (75.4) |
| Moderate to severe | 591 (43.4) | 429 (30.6) | 223 (15.9) | 136 (9.7) | 367 (26.2) | 264 (23.6) | 322 (23.0) | 2,332 (24.6) |
| MAL-ED score, Median (IQR), n (%)<sup>5</sup> | 6 (5, 9) | 6 (5, 8) | 5 (4, 6) | 4 (3, 6) | 6 (4, 7) | 7 (5, 8) | 5 (4, 7) | 6 (4, 7) |
| Non-Severe | 493 (36.2) | 511 (36.5) | 877 (62.7) | 973 (69.5) | 666 (47.6) | 328 (29.4) | 713 (51.0) | 4,561 (48.1) |
| Severe | 868 (63.8) | 889 (63.5) | 522 (37.3) | 427 (30.5) | 734 (52.4) | 789 (70.6) | 686 (49.0) | 4,915 (51.9) |
| *Shigella* mortality score, Median (IQR), n (%)<sup>6</sup> | 2 (0, 3) | 4 (0, 4) | 0 (0, 0) | 0 (0, 2) | 2 (0, 2) | 4 (4, 6) | 0 (0, 0) | 0 (0, 4) |
| Mild | 1,221 (89.7) | 1,174 (83.9) | 1,393 (99.6) | 1,343 (95.9) | 1,273 (90.9) | 585 (52.4) | 1,351 (96.6) | 8,340 (88.0) |
| Moderate | 74 (5.4) | 163 (11.6) | 5 (0.4) | 55 (3.9) | 122 (8.7) | 521 (46.6) | 20 (1.4) | 960 (10.1) |
| Severe | 66 (4.8) | 63 (4.5) | 1 (0.1) | 2 (0.1) | 5 (0.4) | 11 (1.0) | 28 (2.0) | 176 (1.9) |
| ***Shigella*-attributable MAD** |  |  |  |  |  |  |  |  |
| GEMS MSD, n (%)<sup>1</sup> |  |  |  |  |  |  |  |  |
| Less-severe | 226 (58.2) | 60 (34.3) | 138 (79.8) | 182 (75.8) | 213 (57.9) | 24 (10.6) | 235 (59.2) | 1,078 (54.8) |
| Moderate-to-severe | 162 (41.8) | 115 (65.7) | 35 (20.2) | 58 (24.2) | 155 (42.1) | 203 (89.4) | 162 (40.8) | 890 (45.2) |
| Modified Vesikari score (MVS), Median (IQR), n (%)<sup>2</sup> | 8 (7, 12) | 9 (7, 11) | 7 (5, 9) | 6 (5, 8) | 8 (6, 10) | 10 (8, 12) | 7 (6, 9) | 8 (6, 10) |
| Mild | 242 (62.4) | 81 (46.3) | 128 (74.0) | 185 (77.1) | 227 (61.7) | 72 (31.7) | 286 (72.0) | 1,221 (62.0) |
| Moderate | 41 (10.6) | 45 (25.7) | 26 (15.0) | 24 (10.0) | 73 (19.8) | 62 (27.3) | 45 (11.3) | 316 (16.1) |
| Severe | 105 (27.1) | 49 (28.0) | 19 (11.0) | 31 (12.9) | 68 (18.5) | 93 (41.0) | 66 (16.6) | 431 (21.9) |
| Moderate or severe diarrhea by MVS or dysentery, n (%)<sup>3</sup> | 226 (58.2) | 116 (66.3) | 63 (36.4) | 76 (31.7) | 199 (54.1) | 174 (76.7) | 215 (54.2) | 1,069 (54.3) |
| Clark score, Median (IQR), n (%)<sup>4</sup> | 7 (6, 10) | 7 (5, 9) | 5 (4, 7) | 4 (3, 6) | 7 (5, 9) | 7 (5, 9) | 6 (4, 8) | 6 (5, 9) |
| Mild | 235 (60.6) | 127 (72.6) | 149 (86.1) | 215 (89.6) | 262 (71.2) | 158 (69.6) | 316 (79.6) | 1,462 (74.3) |
| Moderate to severe | 153 (39.4) | 48 (27.4) | 24 (13.9) | 25 (10.4) | 106 (28.8) | 69 (30.4) | 81 (20.4) | 506 (25.7) |
| MAL-ED score, Median (IQR), n (%)<sup>5</sup> | 6 (5, 9) | 6 (5, 8) | 5 (4, 6) | 4 (3, 6) | 6 (4, 8) | 7 (6, 9) | 5 (4, 7) | 6 (4, 8) |
| Non-Severe | 158 (40.7) | 56 (32.0) | 116 (67.1) | 164 (68.3) | 154 (41.8) | 49 (21.6) | 217 (54.7) | 914 (46.4) |
| Severe | 230 (59.3) | 119 (68.0) | 57 (32.9) | 76 (31.7) | 214 (58.2) | 178 (78.4) | 180 (45.3) | 1,054 (53.6) |
| *Shigella* mortality score, Median (IQR), n (%)<sup>6</sup> | 2 (0, 3) | 4 (0, 4) | 0 (0, 0) | 0 (0, 2) | 2 (0, 3) | 6 (4, 6) | 0 (0, 0) | 2 (0, 3) |
| Mild | 359 (92.5) | 149 (85.1) | 171 (98.8) | 227 (94.6) | 321 (87.2) | 108 (47.6) | 381 (96.0) | 1,716 (87.2) |
| Moderate | 20 (5.2) | 20 (11.4) | 1 (0.6) | 11 (4.6) | 45 (12.2) | 116 (51.1) | 7 (1.8) | 220 (11.2) |
| Severe | 9 (2.3) | 6 (3.4) | 1 (0.6) | 2 (0.8) | 2 (0.5) | 3 (1.3) | 9 (2.3) | 32 (1.6) |

GEMS: Global Enteric Multicenter Study, MSD: moderate-to-severe diarrhea, MVS: modified Vesikari score, WHO: World Health Organization.
<sup>1</sup> Defined as in Kotloff, Lancet GH, 2019. Moderate-to-severe diarrhea (MSD) defined as presenting to a health facility with diarrhea and severe or some dehydration (by WHO criteria), visible blood in stool, or inpatient admission. Less-severe-diarrhea defined as presenting to a health facility without MSD.
<sup>2</sup> Defined as in PATH Vesikari Clinical Severity Scoring System Manual: Duration of diarrhea: 1-4 days (1 point), 5 days (2 points) ≥6 days (3 points); Max # of stool in 24 hour period: 1-3 (1 point), 4-5 (2 points), ≥6 (3 points); Duration of vomiting: 1 day (1 point), 2 days (2 points), ≥3 days (3 points); max # of vomiting episodes in 24 hour period: 1 (1 point), 2-4 (2 points), ≥5 (3 points); Axillary temperature 36.6-37.9°C (1 point), 38.0-38.4°C (2 points), ≥38.5°C (3 points); dehydration 1-5% (2 points), ≥6% (3 points); treatment: rehydration (1 point), hospitalization (2 points). Classified as mild (0-8 points), moderate (9-10 points), and severe (≥11 points).
<sup>3</sup> Defined as in Pavlinac, Vaccines, 2022 as a MVS of 9+ or presence of visible blood in stool.
<sup>4</sup> Defined as in Clark, JID, 1988. Duration of diarrhea: 1-4 days (1 point), 5-7 days (2 points) >7 days (3 points); Max # of stool in 24 hour period: 2-4 (1 point), 5-7 (2 points), >7 (3 points); Duration of vomiting: 2 days (1 point), 3-5 days (2 points), >5 days (3 points); max # of vomiting episodes in 24 hour period: 1-3 (1 point), 4-6 (2 points), >6 (3 points); Duration of reported fever: 1-2 days (1 point), 3-4 days (2 points), ≥5 days (3 points); Rectal temperature 38-38.2°C (1 point), 38.3-38.7°C (2 points), ≥38.8°C (3 points); behavioral signs: Irritable/less playful (1 point), Lethargic/listless (2 points), Seizures (3 points). Classified as mild (2-8 points) and moderate to severe (≥9 points).
<sup>5</sup> Defined as in Lee, J Pediatr Gastroenterol Nutr., 2016: Duration of diarrhea: 2-4 days (1 point), 5-7 days (2 points), ≥8 days (3 points); Max # of stool in 24 hour period: <5 loose stools/24 hours (1 point), 5-7 loose stools/24 hours (2 points), ≥8 stools/24 hours (3 points); Duration of vomiting: 1 day (1 point), 2 days (2 points), ≥3 days (3 points); duration of reported fever: 1+ days (1 point); Confirmed temperature: ≥37.5°C (confirmed by field worker) (2 points); Dehydration: Some (2 points), severe (3 points). Classified as non-severe (0-5 points) and severe (≥6 points).
<sup>6</sup> Defined as in Pavlinac, CID, 2021. Duration of diarrhea through day of presentation: 1-3 days (0 points), 4-5 days (2 points), ≥6 days (3 points); WHO-defined dehydration categories: severe (8 points), some (4 points), none (0 points); inpatient admission (5 points). Classified as mild (0-5 points), moderate (6-8 points), and severe (≥9 points).

**Figure A1.**
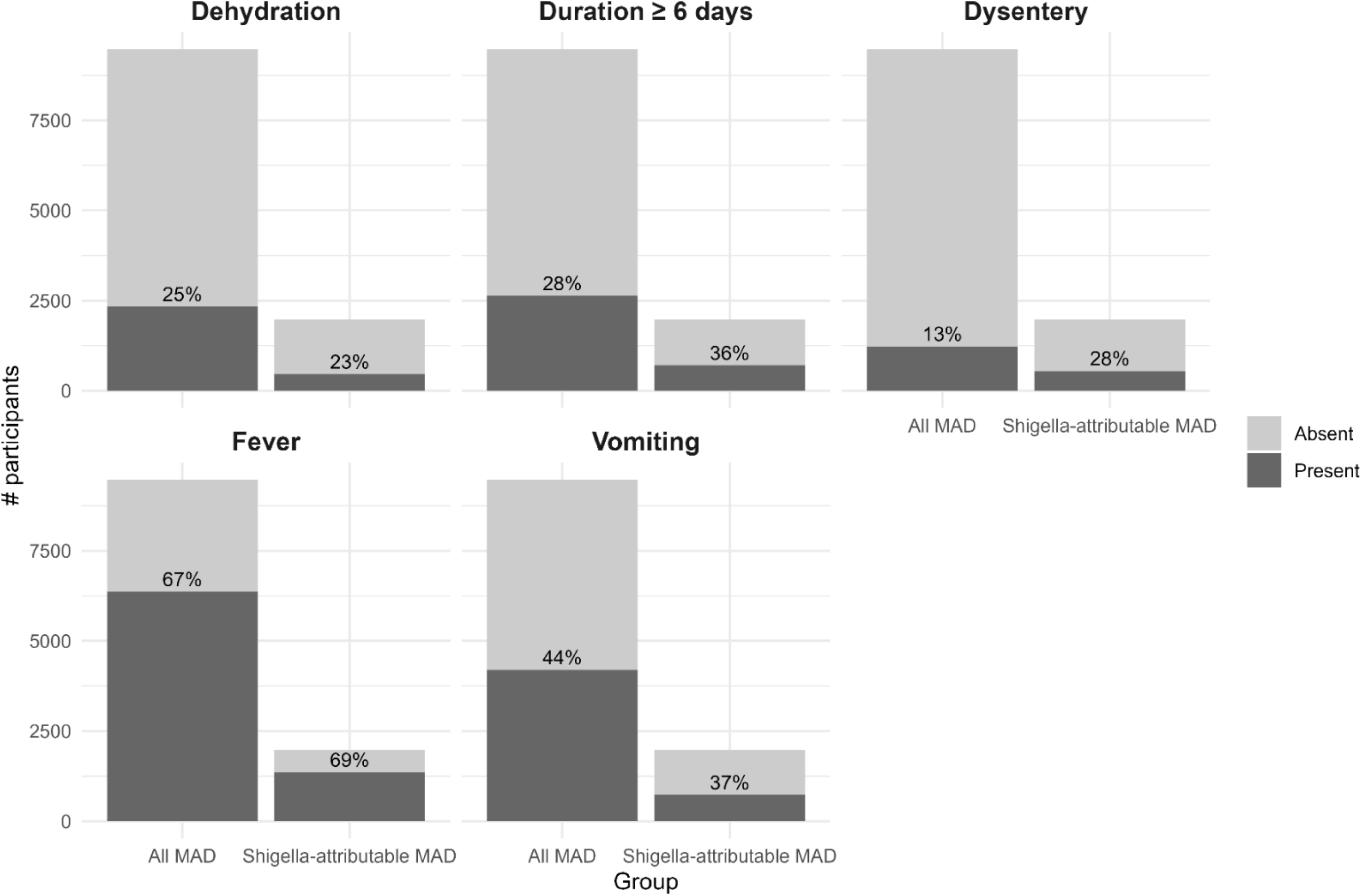
Frequency of severity indicators among all medically-attended diarrhea (MAD) and *Shigella*-attributable MAD. Percentages indicate presence of indicator. Alt text: Bar graph depicting the percentage of children with five severity indicators (dehydration, duration ≥6 days, dysentery, fever, and vomiting) among all medically-attended diarrhea and *Shigella*-attributable medically attended diarrhea.

**Figure A2.**
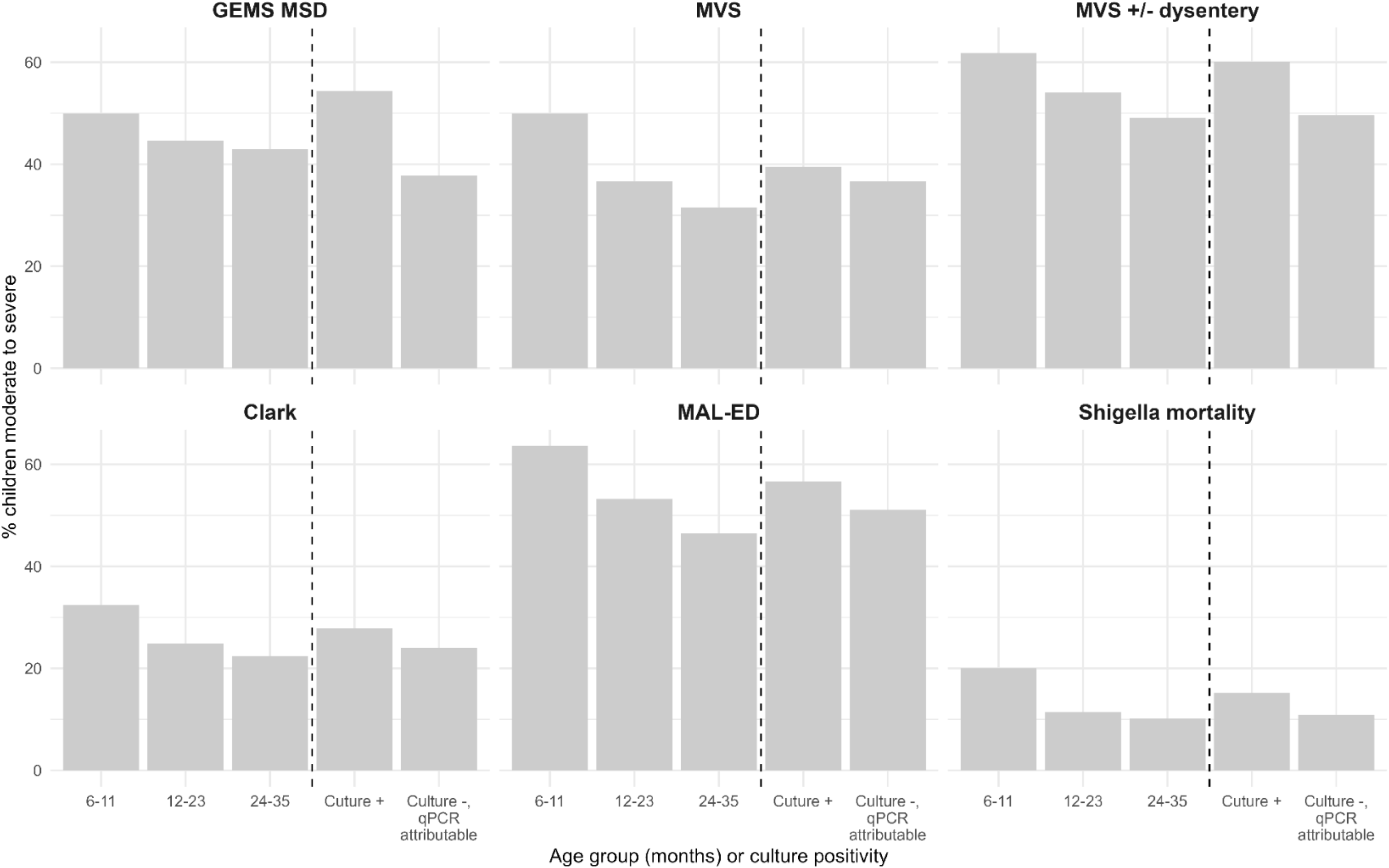
Percentage of children with *Shigella*-attributable medically attended diarrhea classified as having moderate to severe diarrhea by each severity definition by age group and culture positivity. GEMS: Global Enteric Multicenter Study, MSD: moderate-to-severe diarrhea, MVS: modified Vesikari score Scores dichotomized as follows: MVS dichotomized as mild (<9 points) vs. moderate or severe (≥9 points); MVS +/− dysentery dichotomized as mild with no dysentery vs. moderate/severe or dysentery; GEMS MSD dichotomized as less-severe-diarrhea vs. moderate-to-severe-diarrhea; *Shigella* mortality score dichotomized as mild (<6 points) vs. moderate or severe (≥6 points) diarrhea; Clark score dichotomized as mild (<9 points) vs. moderate to severe diarrhea (≥9 points); MAL-ED dichotomized as non-severe (<6 points) vs. severe diarrhea (≥6 points) Alt text: Bar graph depicting the percentage of *Shigella*-attributable medically attended diarrhea classified as moderate or severe by each severity definition by age group (6-11 months, 12-23 months, and 24-25 months) and by culture positivity (culture positive, culture negative qPCR-attributable).

**Table A4.** Association between more severe diarrhea by dichotomized severity definitions and change in length/height-for-age z-score (ΔLAZ/HAZ) from enrollment to 3-month follow-up among medically-attended diarrhea (MAD) and *Shigella*-attributable MAD. Models among all diarrhea cases and stratified by guideline-recommended antibiotic treatment. All models adjusted for baseline LAZ/HAZ, site, and days from enrollment to 3-month follow-up visit.

| Severity definition<sup>1</sup> | Total |  | Guideline-recommended antibiotics |  | No guideline-recommended antibiotics |  |
| --- | --- | --- | --- | --- | --- | --- |
|  | Mean difference (CI) | p-value | Mean difference (CI) | p-value | Mean difference (CI) | p-value |
| **All MAD** | **N = 8767** |  | **N = 3211** |  | **N = 5556** |  |
| GEMS MSD (moderate-to-severe)<sup>2</sup> | 0.00 (-0.02, 0.02) | 0.855 | -0.02 (-0.06, 0.01) | 0.152 | -0.02 (-0.05, 0.01) | 0.165 |
| MVS (moderate or severe)<sup>3</sup> | -0.07 (-0.09, -0.05) | 0.000 | -0.05 (-0.08, -0.03) | 0.000 | -0.08 (-0.11, -0.06) | 0.000 |
| MVS (moderate or severe) and/or dysentery<sup>4</sup> | -0.05 (-0.07, -0.03) | 0.000 | -0.05 (-0.08, -0.02) | 0.001 | -0.07 (-0.10, -0.05) | 0.000 |
| Clark score (moderate to severe)<sup>5</sup> | -0.06 (-0.08, -0.04) | 0.000 | -0.06 (-0.09, -0.03) | 0.000 | -0.07 (-0.09, -0.04) | 0.000 |
| MAL-ED score (severe)<sup>6</sup> | -0.05 (-0.06, -0.03) | 0.000 | -0.05 (-0.07, -0.02) | 0.001 | -0.06 (-0.08, -0.03) | 0.000 |
| *Shigella* mortality score (moderate or severe)<sup>7</sup> | -0.04 (-0.07, -0.01) | 0.013 | -0.07 (-0.12, -0.03) | 0.001 | -0.02 (-0.06, 0.02) | 0.286 |
| ***Shigella*-attributable MAD** | **N = 1832** |  | **N = 984** |  | **N = 848** |  |
| GEMS MSD (moderate-to-severe)<sup>2</sup> | -0.01 (-0.05, 0.02) | 0.471 | -0.05 (-0.10, 0.00) | 0.054 | -0.04 (-0.10, 0.02) | 0.208 |
| MVS (moderate or severe)<sup>3</sup> | -0.07 (-0.10, -0.03) | 0.000 | -0.04 (-0.09, 0.01) | 0.100 | -0.10 (-0.15, -0.04) | 0.000 |
| MVS (moderate or severe) and/or dysentery<sup>4</sup> | -0.05 (-0.09, -0.02) | 0.002 | -0.07 (-0.12, -0.02) | 0.009 | -0.10 (-0.15, -0.05) | 0.000 |
| Clark score (moderate to severe)<sup>5</sup> | -0.03 (-0.07, 0.01) | 0.160 | -0.03 (-0.08, 0.02) | 0.271 | -0.03 (-0.10, 0.03) | 0.319 |
| MAL-ED score (severe)<sup>6</sup> | -0.04 (-0.07, 0.00) | 0.025 | -0.04 (-0.08, 0.01) | 0.121 | -0.05 (-0.10, 0.00) | 0.048 |
| *Shigella* mortality score (moderate or severe)<sup>7</sup> | -0.01 (-0.07, 0.05) | 0.731 | -0.03 (-0.10, 0.05) | 0.485 | 0.00 (-0.10, 0.10) | 0.960 |

CI: 95% confidence interval, GEMS: Global Enteric Multicenter Study, HAZ: Height-for-age z-score, LAZ: Length-for-age z-score, MAL-ED: Malnutrition and Enteric Disease Study, MSD: moderate-to-severe diarrhea, MVS: modified Vesikari score, QIC: quasi information Criterion, WHO: World Health Organization.
<sup>1</sup> Models adjusted for baseline LAZ/HAZ, site, and number of days from enrollment to 3-month follow-up visit.
<sup>2</sup> Defined as in Kotloff, Lancet GH, 2019. Moderate-to-severe diarrhea (MSD) defined as presenting to a health facility with diarrhea and severe or some dehydration (by WHO criteria), visible blood in stool, or inpatient admission.
<sup>3</sup> Defined in PATH Vesikari Clinical Severity Scoring System Manual as a MVS of 9+.
<sup>4</sup> Defined as in Pavlinac, Vaccines, 2022 as a MVS of 9+ or presence of visible blood in stool.
<sup>5</sup> Defined as in Clark, JID, 1988 as a score of 9+.
<sup>6</sup> Defined as in Lee, J Pediatr Gastroenterol Nutr., 2016 as a score of 6+.
<sup>7</sup> Defined as in Pavlinac, CID, 2021 as a score of 6+.

**Table A5.** Diagnostic accuracy of dichotomized severity scores in identifying hospitalization and/or mortality within 14 days of enrollment among medically-attended diarrhea (MAD) and *Shigella*-attributable MAD. Calculated among all diarrhea cases and stratified by guideline-recommended antibiotic treatment.

| Severity definition | Total |  |  |  |  | Guideline-recommended antibiotics |  |  |  |  | No guideline-recommended antibiotics |  |  |  |  |
| --- | --- | --- | --- | --- | --- | --- | --- | --- | --- | --- | --- | --- | --- | --- | --- |
|  | Sensitivity (CI) | Specificity (CI) | Total accuracy (CI) | PPV (CI) | NPV (CI) | Sensitivity (CI) | Specificity (CI) | Total accuracy (CI) | PPV (CI) | NPV (CI) | Sensitivity (CI) | Specificity (CI) | Total accuracy (CI) | PPV (CI) | NPV (CI) |
| **All MAD** | **N = 9476** |  |  |  |  | **N = 3366** |  |  |  |  | **N = 6110** |  |  |  |  |
| GEMS MSD (moderate-to-severe)<sup>1</sup> | 94.2 (91.1, 96.9) | 66.5 (65.6, 67.4) | 67.2 (66.4, 68.1) | 6.8 (6, 7.7) | 99.8 (99.7, 99.9) | 97.9 (95.6, 100) | 61.3 (59.8, 62.8) | 62.9 (61.4, 64.4) | 10.3 (8.7, 11.9) | 99.8 (99.7, 100) | 88.3 (81.7, 94.3) | 69.3 (68.4, 70.3) | 69.6 (68.7, 70.6) | 4.3 (3.4, 5.2) | 99.7 (99.6, 99.9) |
| MVS (moderate or severe)<sup>2</sup> | 88.3 (84.2, 92.4) | 60.1 (59.2, 61.1) | 60.8 (59.9, 61.8) | 5.4 (4.7, 6.2) | 99.5 (99.3, 99.7) | 88.4 (83.3, 93.4) | 60.1 (58.4, 61.8) | 61.3 (59.7, 63) | 9.1 (7.8, 10.8) | 99.1 (98.7, 99.5) | 88.3 (81.3, 94.2) | 60.1 (58.9, 61.3) | 60.5 (59.3, 61.7) | 3.3 (2.6, 4.1) | 99.7 (99.5, 99.9) |
| MVS (moderate or severe) and/or dysentery<sup>3</sup> | 89.6 (85.9, 93.2) | 52.7 (51.7, 53.7) | 53.7 (52.7, 54.6) | 4.7 (4.1, 5.3) | 99.5 (99.3, 99.7) | 89.7 (84.7, 94.4) | 44.3 (42.8, 45.8) | 46.3 (44.8, 47.8) | 6.8 (5.7, 7.9) | 99 (98.4, 99.4) | 89.4 (82.5, 95.1) | 57.2 (56.1, 58.4) | 57.7 (56.6, 58.9) | 3.2 (2.5, 3.8) | 99.7 (99.5, 99.9) |
| Clark score (moderate to severe)<sup>4</sup> | 70.8 (65.5, 76.3) | 76.6 (75.7, 77.4) | 76.4 (75.6, 77.3) | 7.3 (6.3, 8.3) | 99 (98.8, 99.2) | 74.7 (67.5, 81.4) | 71.7 (70.2, 73.2) | 71.8 (70.4, 73.3) | 10.7 (8.8, 12.5) | 98.4 (97.9, 98.9) | 64.9 (55.7, 74.2) | 79.2 (78.2, 80.2) | 79 (78, 80) | 4.7 (3.6, 5.8) | 99.3 (99.1, 99.5) |
| MAL-ED score<sup>5</sup> | 90 (86.1, 93.7) | 49.1 (48.1, 50.1) | 50.2 (49.2, 51.1) | 4.4 (3.8, 5) | 99.5 (99.3, 99.7) | 91.1 (85.9, 95.2) | 47 (45.2, 48.8) | 49 (47.2, 50.7) | 7.2 (6, 8.5) | 99.1 (98.6, 99.5) | 88.3 (81.8, 94.4) | 50.2 (49, 51.4) | 50.8 (49.6, 52) | 2.7 (2.2, 3.2) | 99.6 (99.4, 99.8) |
| *Shigella* mortality score (moderate or severe)<sup>6</sup> | 72.9 (67, 78.8) | 89.6 (89, 90.2) | 89.2 (88.6, 89.8) | 15.4 (13.2, 17.7) | 99.2 (99, 99.4) | 77.4 (70.3, 84.2) | 91.7 (90.8, 92.6) | 91.1 (90.2, 92) | 29.7 (25.4, 34.5) | 98.9 (98.5, 99.2) | 66 (56.3, 76) | 88.5 (87.7, 89.2) | 88.1 (87.4, 88.9) | 8.2 (6.5, 10.1) | 99.4 (99.2, 99.6) |
| ***Shigella*-attributable MAD** | **N = 1968** |  |  |  |  | **N = 1040** |  |  |  |  | **N = 928** |  |  |  |  |
| GEMS MSD (moderate-to-severe)<sup>1</sup> | 93.6 (85.4, 100) | 56 (53.8, 58) | 56.9 (54.8, 58.8) | 4.9 (3.6, 6.4) | 99.7 (99.4, 100) | 94.4 (86.1, 100) | 45.8 (42.9, 48.5) | 47.5 (44.6, 50.1) | 5.9 (4.1, 7.7) | 99.6 (98.9, 100) | 90.9 (66.7, 100) | 67.1 (64.6, 69.6) | 67.3 (65, 69.8) | 3.2 (1.5, 5.2) | 99.8 (99.5, 100) |
| MVS (moderate or severe)<sup>2</sup> | 80.9 (69.7, 91.7) | 63.1 (61, 65) | 63.5 (61.5, 65.5) | 5.1 (3.6, 6.6) | 99.3 (98.8, 99.7) | 83.3 (70, 94.4) | 64.1 (61.4, 67.1) | 64.8 (62.2, 67.6) | 7.7 (5.1, 10.4) | 99.1 (98.3, 99.7) | 72.7 (40, 100) | 61.9 (58.9, 64.8) | 62.1 (59.1, 65) | 2.2 (0.8, 3.8) | 99.5 (98.8, 100) |
| MVS (moderate or severe) and/or dysentery<sup>3</sup> | 85.1 (74.3, 94.5) | 46.4 (44.1, 48.7) | 47.4 (45.2, 49.6) | 3.7 (2.7, 4.9) | 99.2 (98.6, 99.8) | 88.9 (78, 97.6) | 37.1 (34.3, 39.7) | 38.8 (36.2, 41.6) | 4.8 (3.3, 6.4) | 98.9 (97.9, 99.8) | 72.7 (44.4, 100) | 56.7 (53.6, 59.8) | 56.9 (53.8, 60) | 2 (0.7, 3.4) | 99.4 (98.8, 100) |
| Clark score (moderate to severe)<sup>4</sup> | 66 (52.6, 79.4) | 75.3 (73.4, 77.1) | 75.1 (73.1, 77) | 6.1 (4.2, 8.1) | 98.9 (98.3, 99.4) | 72.2 (57.1, 86.7) | 72 (69.4, 74.7) | 72 (69.3, 74.7) | 8.5 (5.5, 11.6) | 98.6 (97.8, 99.4) | 45.5 (16.7, 80) | 78.8 (76.2, 81.4) | 78.4 (75.7, 81) | 2.5 (0.5, 5) | 99.2 (98.5, 99.7) |
| MAL-ED score<sup>5</sup> | 87.2 (77.8, 95.7) | 47.3 (45.1, 49.2) | 48.2 (46, 50.2) | 3.9 (2.8, 5) | 99.3 (98.8, 99.8) | 86.1 (73.3, 97.2) | 45.3 (42.3, 48) | 46.7 (43.8, 49.4) | 5.3 (3.5, 7.1) | 98.9 (97.8, 99.8) | 90.9 (66.7, 100) | 49.4 (46.3, 52.5) | 49.9 (46.8, 53) | 2.1 (0.9, 3.4) | 99.8 (99.3, 100) |
| *Shigella* mortality score (moderate or severe)<sup>6</sup> | 66 (51.2, 79) | 88.5 (87.3, 89.7) | 88 (86.7, 89.2) | 12.3 (8.6, 16.4) | 99.1 (98.5, 99.5) | 63.9 (47.7, 80) | 89.3 (87.6, 90.8) | 88.5 (86.7, 90) | 17.7 (11.6, 23.9) | 98.6 (97.7, 99.3) | 72.7 (45.4, 100) | 87.6 (85.8, 89.4) | 87.4 (85.5, 89.3) | 6.6 (2.8, 10.9) | 99.6 (99.1, 100) |

CI: 95% confidence interval, GEMS: Global Enteric Multicenter Study, HAZ: Height-for-age z-score, LAZ: Length-for-age z-score, MAL-ED: Malnutrition and Enteric Disease Study, MSD: moderate-to-severe diarrhea, MVS: modified Vesikari score, WHO: World Health Organization.
<sup>1</sup> Defined as in Kotloff, Lancet GH, 2019. Moderate-to-severe diarrhea (MSD) defined as presenting to a health facility with diarrhea and severe or some dehydration (by WHO criteria), visible blood in stool, or inpatient admission.
<sup>2</sup> Defined as in PATH Vesikari Clinical Severity Scoring System Manual as a MVS of 9+.
<sup>3</sup> Defined as in Pavlinac, Vaccines, 2022 as a MVS of 9+ or presence of visible blood in stool.
<sup>4</sup> Defined as in Clark, JID, 1988 as a score of 9+.
<sup>5</sup> Defined as in Lee, J Pediatr Gastroenterol Nutr., 2016 as a score of 6+.
<sup>6</sup> Defined as in Pavlinac, CID, 2021 as a score of 6+.

**Figure A3.**
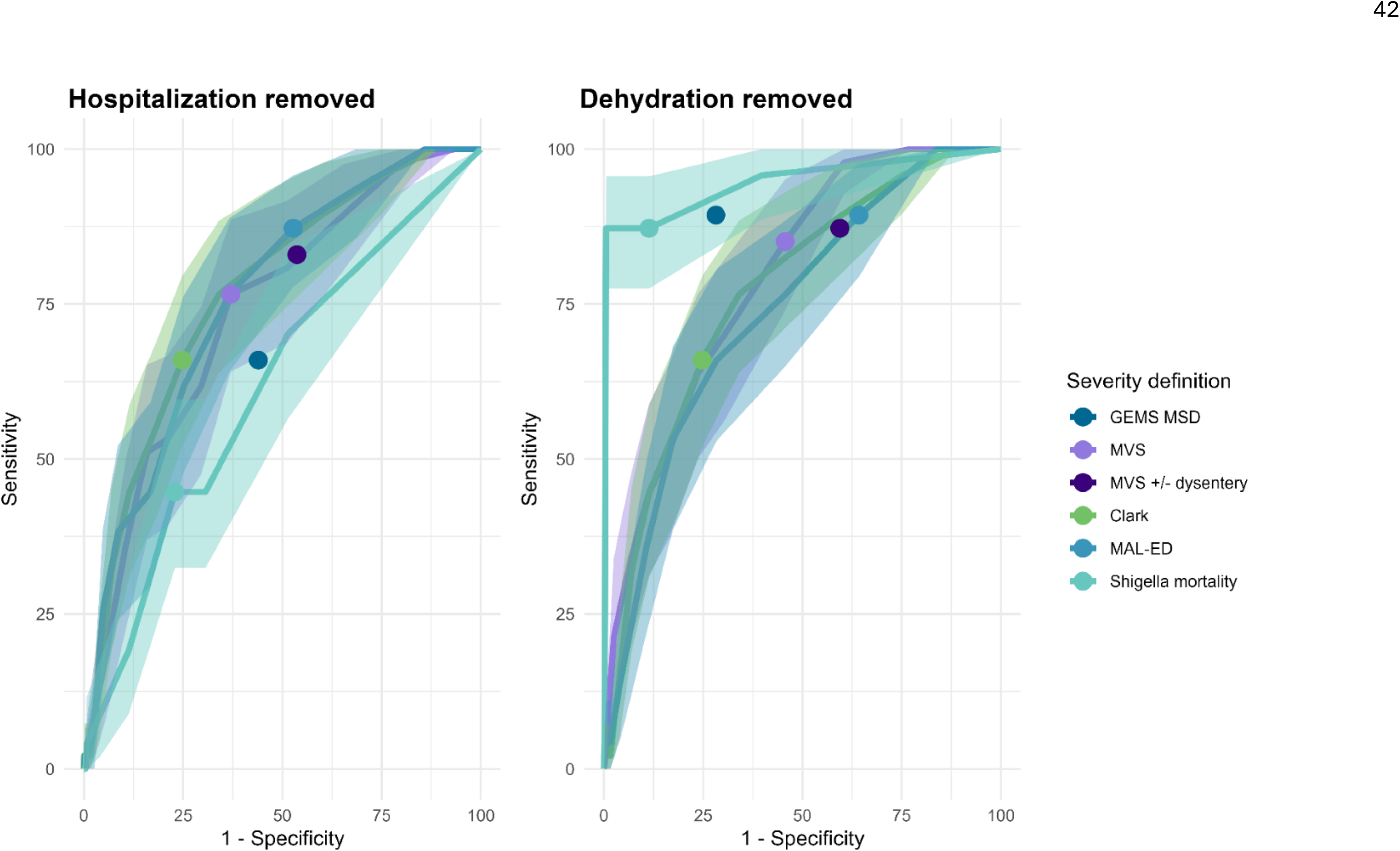
Receiver operating characteristic (ROC) curves of four continuous severity scores with the dehydration and hospitalization components removed, respectively, on the outcome of mortality or hospitalization within 14 days of enrollment among children with *Shigella*-attributable medically-attended diarrhea. Shaded bands represent 95% confidence intervals. Plotted points represent sensitivity and specificity of threshold used for dichotomizing four continuous scores, as well as sensitivity and specificity of two non-numeric severity definitions. GEMS: Global Enteric Multicenter Study, MSD: moderate-to-severe diarrhea, MVS: modified Vesikari score Scores dichotomized as follows: MVS dichotomized as mild (<9 points) vs. moderate or severe (≥9 points); MVS +/− dysentery dichotomized as mild with no dysentery vs. moderate/severe or dysentery; GEMS MSD dichotomized as less-severe-diarrhea vs. moderate-to-severe-diarrhea; *Shigella* mortality score dichotomized as mild (<6 points) vs. moderate or severe (≥6 points) diarrhea; Clark score dichotomized as mild (<9 points) vs. moderate to severe diarrhea (≥9 points); MAL-ED dichotomized as non-severe (<6 points) vs. severe diarrhea (≥6 points) Alt text: Two receiver operating characteristic curves of four continuous severity scores on hospitalization or mortality within 14 days of enrollment among children with *Shigella*-attributable medically-attended diarrhea. In the left plot the hospitalization component has been removed from the scores, and *Shigella* mortality score has lower area under the curve compared to the other three scores (MVS, Clark score, and MAL-ED score), which perform similarly. In the right plot the dehydration component has been removed from the scores and Shigella mortality score has a higher area under the curve than the other scores, which perform similarly.

